# Epidemiological, microbiological, and genomic risk factors for healthcare-associated Carbapenemase producing Enterobacterales (CPE) outbreaks: A systematic review

**DOI:** 10.64898/2025.12.23.25342705

**Authors:** Dorottya Nagy, Alice Baker, Caitlin Barton-Sargeant, Jun Jonathan Yang, William Matlock, Susan Hopkins, Ann Sarah Walker, Alice Ledda, Julie Robotham, Samuel Lipworth, Nicole Stoesser

**Author notes:** **Corresponding author and reprint requests: Name**: Dorottya Nagy, **Correspondence address:** Modernising Medical Microbiology Unit, Experimental Medicine Division, Level 5, Room 5800, Nuffield Department of Medicine, John Radcliffe Hospital, Headley Way, Headington, OX3 9DU, UK, **Email:**. Joint senior authors (Samuel Lipworth, Nicole Stoesser).

## Abstract

**Background:** Healthcare-associated carbapenemase-producing Enterobacterales (CPE) outbreaks are a major healthcare challenge. Epidemiological studies have identified patient-level risk factors for CPE transmission, and genomic studies have highlighted high-risk’ lineages or mobile genetic elements (MGEs); however, a unified dissemination risk-prediction framework is lacking.

**Objectives:** To synthesise available data on epidemiological, microbiological and genomic risk factors to quantify healthcare-associated CPE outbreak potential.

**Methods:** *Data:* Six bibliographic databases and other sources were searched (‘carbapenemase’ AND ‘outbreak’ AND ‘MGE’; ≤31/01/24). Data were extracted on primary (patients infected/colonised) and secondary (outbreak duration/resolution, mortality) outcomes, and risk/protective factors including epidemiological, microbiological/genomic and infection control measures.

*Study eligibility:* Studies reporting healthcare-associated CPE outbreaks involving MGE-associated IMP/KPC/NDM/OXA-48-like/VIM carbapenemases confirmed by whole-genome sequencing.

*Study quality:* Reporting quality was assessed against the ORION checklist (random subset).

*Data synthesis:* After descriptive summaries, multivariable linear mixed effect modelling was used to estimate associations between risk/protective factors and outbreak size.

**Results:** 179 records (272 outbreaks) were included from 3,188 screened (41 countries, 2004-2023), affecting median 10 patients (IQR=5-27, range=2-223), and lasting 12 months (IQR=5-30, range=1 day-16 years). Data on outbreak size (primary outcome) was 99.6% complete (271/272) but more limited for secondary outcomes (29-97% complete) and risk/protective factors (70/91 factors had ≥10% missingness). 39% (107/272) of outbreaks involved MGE-mediated transmission, which is a potential underestimate as 66% (104/157) of reports used clonal outbreak definitions. The involvement of more institutions (adjusted relative outbreak size: 1.10 per institution [95% CI: 1.04-1.16];*p=0.001*), and more Enterobacterales sequence types (1.04 per sequence type [1.01-1.08];*p=0.011*), were associated with larger outbreaks. Reporting quality assessment (n=98 studies) revealed adequate reporting on median 11/19 relevant ORION items (IQR=8-13; range=1-18).

**Conclusions:** Heterogenous/incomplete reporting of CPE outbreaks precludes integrated risk evaluation based on epidemiological, microbiological, and genomic factors. Systematic sampling, sequencing and epidemiological metadata reporting may strengthen data quality for quantifying healthcare-associated CPE dissemination risk.

## Introduction

Carbapenemase-producing Enterobacterales (CPE) are WHO ‘critical priority’ pathogens(1) and targeted by several National Action Plans(2–5). Carbapenemases are particularly concerning in healthcare settings, conferring resistance to ‘reserve-list’ antibiotics(6, 7), and are associated with higher mortality and healthcare costs(8, 9). Furthermore, the genes encoding them have the capacity for undetected transmission amongst asymptomatically colonised patients, healthcare workers and environmental reservoirs(10–14).

The ‘big five’ carbapenemase gene families (*bla*_OXA-48-like_, *bla*_KPC_, *bla*_NDM_, *bla*_VIM_, *bla*_IMP_) are the most clinically relevant globally(15–17), and are commonly encoded on mobile genetic elements (MGEs, i.e. plasmids, transposons/insertion sequences [ISs] or integrons)(7, 13, 17–19), facilitating horizontal gene transfer (HGT). HGT may occur across bacterial lineages and/or species, potentially delaying outbreak detection where conventional lineage-based outbreak definitions are applied. The increased accessibility of long-read and hybrid whole genome sequencing (WGS) may enable tracking of AMR gene (ARG)-associated MGEs including plasmids with greater resolution(20–22), potentially contributing to a paradigm shift in surveillance.

While genomic studies have implicated ‘high-risk’ lineages and MGEs/plasmids(18, 23), and epidemiological studies have evaluated patient-level risk factors for CPE transmission(24, 25), a joint epidemiological, microbiological and genomic framework quantifying dissemination risk for emerging CPE-lineages or plasmids in healthcare is lacking. Clear evidence to support the allocation of resources in the early stages of outbreak investigations and infection prevention and control (IPC) at both institutional and public health levels is thus limited.

This systematic review aimed to synthesise available data on epidemiological, microbiological and genomic risk factors and IPC measures for healthcare-associated CPE outbreaks mediated by the ‘big five’ carbapenemases through an ecological analysis, and construct a unified framework to predict healthcare-associated CPE dissemination risk.

## Methods

### Design

A systematic review of publicly available reports of healthcare-associated CPE outbreaks was conducted. Included studies were synthesised using an ecological study, with each outbreak treated as an equally-weighted data point.

### Inclusion & exclusion criteria Patients

Studies on patients of all ages attending healthcare facilities, colonised/infected with CPE in outbreak/epidemic situations attributed to one of the ‘big five’ carbapenemase gene families, were included. Studies lacking any epidemiological metadata (date, location, disease status or other patient characteristics) were excluded.

### Exposure/Intervention

Risk/protective factors (exposures) for outbreaks were recorded from each study under four domains (study features, epidemiological aspects, microbiological/genomic factors, infection control measures; Table 1).

**Table 1.** Outbreak features extracted under four domains: study features, epidemiological aspects, microbiological/genomic factors and infection control measures.

| Study features | Epidemiological aspects | Microbiological / genomic factors | Infection control measures |
| --- | --- | --- | --- |
| • First author<br>• Journal<br>• Publication year<br>• Publication type<br>• Study design<br>• Funding<br>• Conflict of interest<br>• Number of outbreaks reported in study<br>• ORION (Outbreak Reports and Intervention Studies Of Nosocomial infection checklist) quality assessment and risk of bias checklist | • Start year (Year of detection)<br>• Continent<br>• Country<br>• Number/type/capacity of institutions involved<br>• Number/type of wards involved<br>• Endemic or epidemic setting (evidence of background prevalence of epidemic carbapenemase gene in hospital or hospital catchment population)<br>• Method of outbreak detection<br>○ Clinical/ screening/ retrospective<br>• Transmission mechanisms involved (non-mutually exclusive)<br>○ Clonal<br>○ Plasmid<br>○ transposon/IS/integron<br>• Outbreak definition criteria (non-mutually exclusive)<br>○ Epidemiological<br>○ Clonal<br>○ Plasmid<br>○ small MGE<br>○ Gene-based | • Number and identity of carbapenemase-producing species and sequence types (STs)<br>• Involvement of non-Enterobacterales species<br>• Carbapenemase gene involved<br>○ Gene family<br>○ Specific variant(s)<br>○ Number of different alleles/variants<br>• Number of phenotypic resistances per isolate<br>• Number of ARGs per isolate<br>• Number of virulence genes per isolate<br>• Other MGEs involved per isolate<br>○ Number<br>○ Type (integron/ transposon/ IS)<br>○ Specific MGE identity<br>• Plasmids<br>○ Number of carbapenemase-associated plasmids<br>○ Number of incompatibility (Inc) types<br>○ Identity of main epidemic plasmid Inc type<br>○ Identity/name of main plasmid backbone<br>○ Main plasmid size<br>○ Number of ARGs on main plasmid<br>○ Number of MGEs on main plasmid<br>○ Main plasmid conjugation ability | • Patient control measures<br>○ Enhanced admission screening<br>○ Patient cohorting<br>○ Contact precautions<br>○ Screening during hospitalisation<br>○ Contact tracing<br>○ Other patient measures<br>• Healthcare worker (HCW) control measures<br>○ HWC screening<br>○ HCW education<br>○ Hand hygiene enhancement<br>○ HCW cohorting<br>○ Outbreak response team establishment<br>○ Other HCW measures including antibiotic use change and personal protective equipment<br>• Environmental control measures<br>○ Environmental screening<br>○ Environmental cleaning<br>○ Replacement of equipment/ furnishings<br>○ Wards closure<br>○ Other environmental measures |

### Context

Eligible contexts were CPE outbreak/epidemic situations where reports included at least one instance of within-institution transmission of one of the ‘big five’ carbapenemase genes (*bla*_OXA-48-like_, *bla*_KPC_, *bla*_NDM_, *bla*_VIM_, or *bla*_IMP_). Carbapenemase genes had to be associated with MGEs (i.e. on plasmids/transposons/integrons; near ISs), confirmed by WGS on ≥1 outbreak isolate, but the outbreak could be clonal or MGE-mediated or both. Where available, the included study authors’ own outbreak definition was used.

### Outcome

The primary outcome was outbreak size (number of patients infected/colonised per outbreak). Secondary outcome measures were outbreak duration, outbreak ‘resolution status’ (i.e. binary reversion/non-reversion of incidence of infection/colonisation to baseline/zero, or establishment of institutional endemicity), number/proportion of patients infected, colonised and deceased (authors’ mortality definition if all-cause mortality unavailable). The study authors’ own definitions of ‘outbreak-associated’ and ‘duration’ were used.

### Study eligibility criteria

Included study types were primary peer-reviewed studies/publications, pre-prints and conference abstracts describing CPE outbreaks in humans (infection/colonisation) in healthcare-associated (i.e. hospital and long-term healthcare) settings. Eligible study designs included outbreak reports, surveillance reports, case series, interrupted time series, controlled before-after studies (quasi-experimental studies), case-control, cohort, cross-sectional, and ecological studies. Reference lists for review articles were also considered. Isolated case reports, retracted publications, comments, errata, and letters were excluded.

### Search strategy

Electronic literature searches were performed using piloted key-word searches and subject headings combining the concepts (carbapenemase) AND (outbreak) AND (plasmid OR other MGEs), up to 31/01/24, without language restrictions (Table S1). Six bibliographic databases/search engines were searched (MEDLINE, EMBASE, Global Health, Scopus, Web of Science, Global Index Medicus). Two preprint databases (BioRxiv, MedRxiv), three conference proceedings (ECCMID [2012-2023], ASM Microbe [2020-2023], IDSA IDWeek [2014-2024]), and three disease surveillance registers (The Worldwide Database for Nosocomial Outbreaks, US CDC, European CDC) were also searched. Expert consultation and backwards citation searching were undertaken (Table S1). The review was registered in PROSPERO (<u>CRD42024505048</u>).

### Study reporting quality assessments

A randomly selected subset of 100 studies was selected for evaluation using the Outbreak Reports and Intervention Studies of Nosocomial Infection (ORION) checklist(26, 27). 19/22 ORION checklist items were considered relevant to this review (item 12 [‘economic analyses’], item 14 [‘sample size’], and item 16 [‘recruitment’] were considered not relevant/not applicable). Checklist item 13 (‘potential threats to internal validity’) was interpreted as reporting factors that may affect case detection during the outbreak period (e.g. changes in testing/screening protocols, laboratory workflows, admission/patient management, antimicrobial stewardship, staffing, hand-hygiene/other IPC measures, and seasonality). Study reporting quality was not used as an exclusion criterion.

### Data

Records were de-duplicated, screened, and data extracted using the systematic review management software Covidence. Statistical analyses were performed in R(28)(v4.4.1). The cleaned dataset and analysis scripts are available online (https://figshare.com/articles/dataset/Extraction_data_and_R_analysis_script_for_CPE_systematic_review/30148717).

### Data screening

Following de-duplication, two independent reviewers (DN and AB) screened all studies for inclusion based on title/abstract, with conflicts adjudicated by a third reviewer (NS/SL). Full-text screening was conducted by the same two reviewers, and conflicts resolved by discussion (Fig.1).

**Figure 1:**
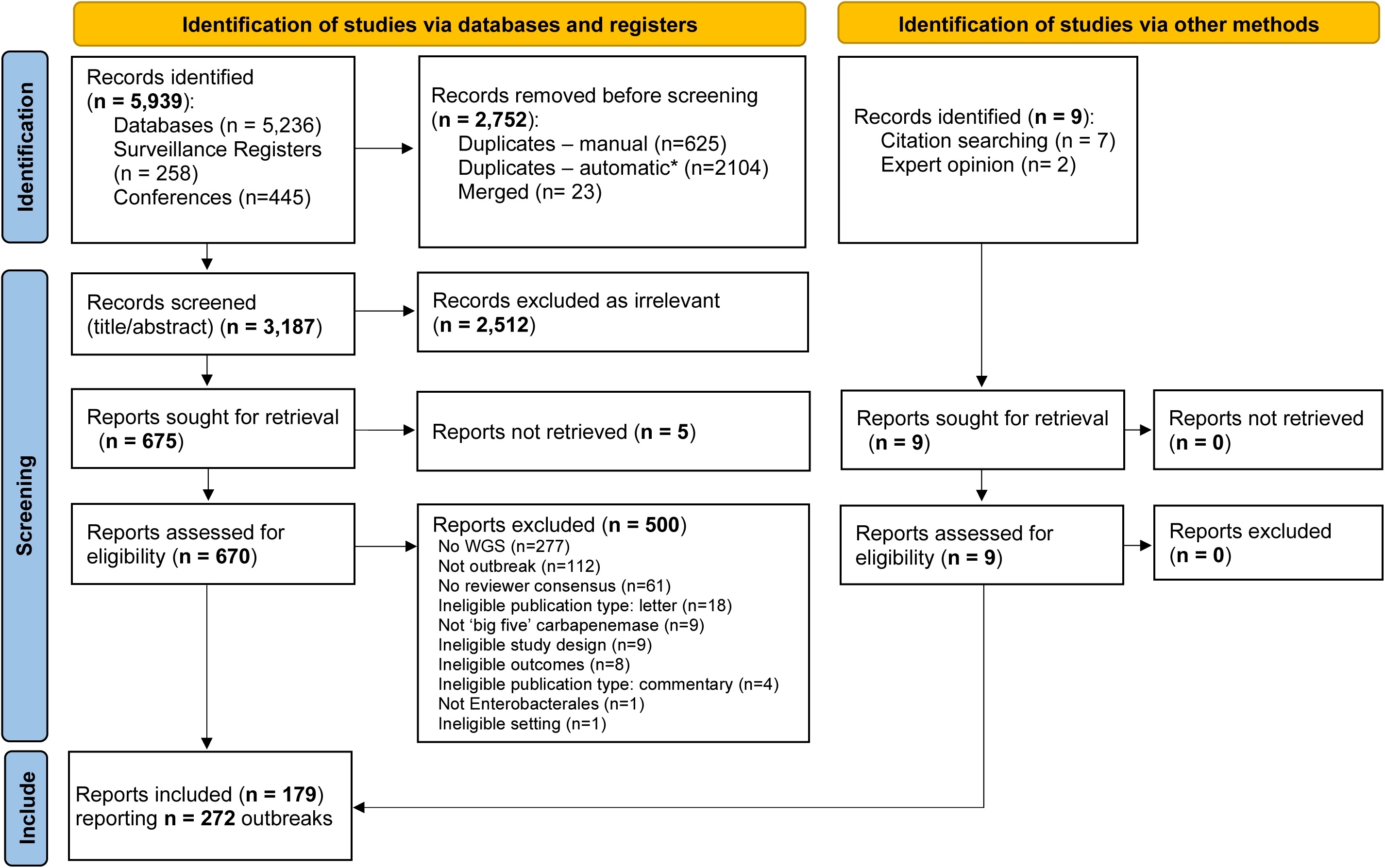
PRISMA diagram showing number of records retrieved, excluded at each stage of screening, and included in final analysis, following standard PRISMA structure. *Automatic record deduplication performed by the Covidence software. Abbreviations: WGS – Whole Genome Sequencing.

### Data extraction

Data was extracted by a single reviewer for all records (DN), and by a second independent reviewer for a randomly selected 20% of records (AB, CSB or JJY), following Cochrane guidelines(29). Conflicts were resolved by discussion.

### Analysis

Inter-rater agreement was assessed using Cohen’s Kappa statistic. Extracted variables reported in ≥12 records were summarised descriptively. Continuous/ordinal variables were summarised with medians/IQRs, and categorical variables with percentages. Continuous variables with outliers (>1.5xIQR below the 25^th^ or above the 75^th^ percentiles) were truncated at the 5^th^ and 95^th^ percentiles to reduce outlier influence.

Univariable linear regression (for log_10_[total cases per outbreak], approximately normally distributed) was used to calculate unadjusted coefficients for each risk/protective factor reported in ≥12 records. A threshold of 12 records was chosen as variables reported in fewer records could not be fitted during univariable linear regression due to data sparsity. A core multivariable linear mixed effect model for log_10_(total cases per outbreak) was fitted using backwards selection, after assessing for multicollinearity, to estimate adjusted effect sizes of risk/protective factors with ≥90% complete data, with study as the random effect to account for non-independence of multiple outbreaks reported per study (‘all-outbreaks’ final model). Variables with *p<0.15* were retained during backwards selection. In cases of high autocorrelation (ρ>0.7), separate models were constructed for each variable, keeping other variables identical, and the model with the lowest Akaike Information Criterion (AIC) selected. All coefficients and confidence intervals (CIs) were back transformed to the original scale for interpretation as a multiplicative fold-change in outbreak size.

For potential risk/protective factors (with ≥12 records) not in the core model (<90% complete or >90% complete and not selected), each was added individually to the core model to obtain an adjusted effect estimate. Sensitivity analysis repeated regression analyses for outbreaks with ≥10 patients (‘10-or-more-patients’ outbreak model) to assess for a potential ‘ceiling effect’ of very small outbreaks (i.e. an outbreak of two patients cannot involve 10 institutions). P-values were assessed with a Wald test, using Satterthwaite’s method with a type 3 ANOVA table for categorical variables overall(30).

## Results

After screening 3,188 non-duplicate records, 179 records were included, reporting 272 outbreaks (Fig.1; Table 2). Inter-rater agreement was acceptable during abstract/title (κ=0.65) and good at full-text (κ=0.81) screening. The 179 studies, published 2013-2024, were predominantly peer-reviewed (157/179 [88%]) outbreak reports (115/179 [64%]), reporting 1-9 outbreaks per study (Tables S2-3).

**Table 2:** Summary of outbreak outcomes reported for 272 outbreaks from 179 included studies.

| Outbreak outcome | Missing number (%) | Number (%) outbreaks with data available | Median (IQR) or number (%) of outbreaks | Range |
| --- | --- | --- | --- | --- |
| **Patients affected (infected/colonised; n=6,175)** |  |  |  |  |
| Number of patients affected per outbreak | 1 (0.4%) | 271 (99.6%) | 10 (5-26) | 2-223 |
| **Colonised (n=1,895)** |  |  |  |  |
| Number of patients colonised per outbreak | 100 (36.8%) | 172 (63.2%) | 3 (0-11) | 0-131 |
| Proportion colonised |  |  | 0.40 (0.00-0.79) | 0.00-1.00 |
| **Infected (n=2,082)** |  |  |  |  |
| Number of patients infected per outbreak | 98 (36.0%) | 174 (64.0%) | 6 (2-14) | 0-100 |
| Proportion infected |  |  | 0.54 (0.22-1.00) | 0.00-1.00 |
| **Deceased (n=415)** |  |  |  |  |
| Number of patients deceased per outbreak | 192 (70.6%) | 80 (29.4%) | 3 (1-6) | 0-35 |
| Proportion deceased |  |  | 0.30 (0.12-0.47) | 0.00-1.00 |
| **Duration (months)** | 9 (3.3%) | 263 (96.7%) | 11.6 (4.9-29.9) | 0-191.6 |
| **Outbreak 'resolution'** |  |  |  |  |
| Yes | 123 (45.2%) | 149 (54.8%) | 82 (55.0%) | - |

The 272 outbreaks occurred from 2004-2023, in 41 countries across 6 continents. All top 10 reporting countries were in Europe, North America or Asia (Fig.2). Most outbreaks were reported by China (38 [14%]), USA (25 [9%]), and UK (23 [8%])(Table S4). Globally, the most commonly reported outbreak-associated carbapenemase gene families were *bla*_KPC_ (83/272 [31%]), *bla*_NDM_ (78/272 [29%]) and *bla*_OXA-48-like_ (51/272 [19%]), but with regional variation (Fig.2; Table S5).

**Figure 2:**
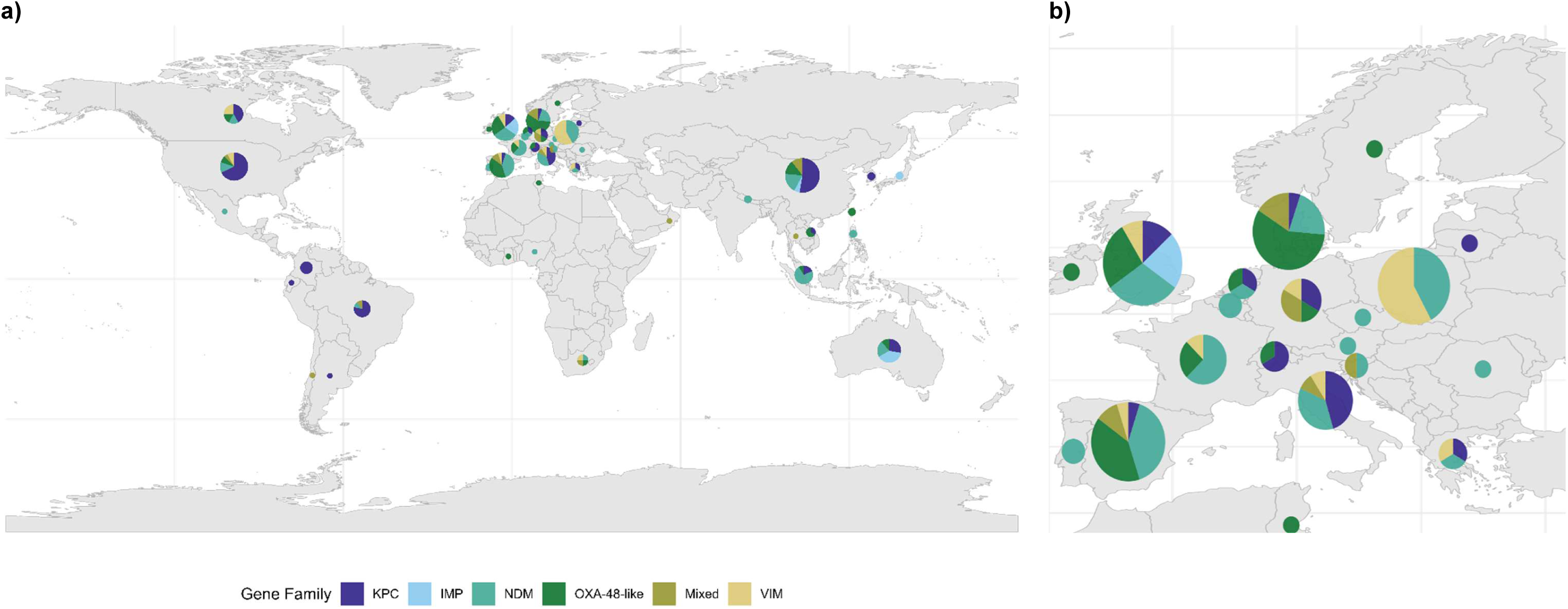
a) World map showing the distribution of hospital outbreak-associated carbapenemase gene families by country. b) Close-up map of Europe. Pie chart size is proportional to the number of outbreaks included from that country.

Reporting of the primary outbreak outcome (number of patients infected/colonised) was the most complete (271/272 [99.6%]). Reporting of secondary outcomes was variable, with 96.7% (263/272) of studies reporting duration, 64.0% (174/272), 63.2% (172/272) and 29.4% (80/272) reporting infection, colonisation and mortality, respectively, and 54.8% (149/272) reporting resolution status (Table 2). Regression was thus performed for the primary outcome only, including the 271/272 outbreaks with complete outbreak size data.

Risk/protective factors reported in ≥12 outbreaks were included in quantitative analyses (93 variables: 8 study features, 35 epidemiological, 29 microbiological/genomic, and 21 infection control measures)(Tables S3-6). Of these, 21 had <10% missing data and were considered for backwards selection in the final core model (Table 3; Fig.3c; Fig.S1).

**Table 3:** Descriptive summary, univariable and multivariable associations between all variables in the final adjusted multivariable mixed effect linear regression model for log10(number of patients affected per outbreak) (‘all-outbreaks’ model). Descriptive summaries are provided considering the dataset of 271 outbreaks with complete outcome data (out of 272 total outbreaks included in this review). Univariable associations are calculated using the outbreaks with complete data for each variable (shown in the 3^rd^ column from the left). Multivariable associations are calculated from a dataset of 256 outbreaks with complete data on all variables included in the final model. Estimates and 95% confidence intervals (CIs) are reported on the original (non-log_10_) scale, interpreted as relative changes in outbreak size (fold-change in size). The final core model had a marginal R^2^ (proportion of the variance explained by fixed effects) of 0.367, conditional R^2^ (proportion of variance explained by fixed and random effects) of 0.499, AIC 260.1, and BIC of 292.0.

| Variable | Category | Number (%)<br>missing | Number (%)<br>of outbreaks<br>with data<br>available | Median (IQR)<br>or<br>Number (%) | Range | Univariable<br>Fold change<br>(95% CIs) | p-value<sup>†</sup> | Multivariable<br>Fold change<br>(95% CIs)<br>(N=256) | p-value<sup>†</sup> |
| --- | --- | --- | --- | --- | --- | --- | --- | --- | --- |
| **Clonal transmission** |  |  |  |  |  |  |  |  |  |
|  | absent | - | - | 18 (6.6%) | - | reference | - | - | - |
|  | present | 0 (0%) | 271 (100%) | 253 (93.4%) | - | 1.20 (0.69-2.08) | 0.518 | 1.82 (1.15-2.88) | **0.011** |
| **Number of institutions\*** |  | 12 (4.4%) | 259 (95.6%) | 1 (1-2) | 1-9 | 1.20 (1.11-1.28) | **<0.001** | 1.18 (1.11-1.25) | **<0.001** |
| **Number of Enterobacterales STs\*** |  | 0 (0%) | 271 (100%) | 1 (1-3) | 1-10 | 1.16 (1.11-1.20) | **<0.001** | 1.14 (1.10-1.19) | **<0.001** |
| **Publication year\*** |  | 0 (0%) | 271 (100%) | 2021 (2019-2022) | 2016-2023 | 1.04 (0.97-1.11) | 0.248 | 1.11 (1.03-1.18) | **0.004** |
| **Start year\*** |  | 3 (1.1%) | 268 (98.9%) | 2016 (2014-2018) | 2010-2021 | 0.94 (0.90-0.98) | **0.007** | 0.92 (0.88-0.97) | **0.001** |
| **Number of outbreaks in study** |  | 0 (0%) | 271 (100%) | 1 (1-4) | 1-9 | 0.86 (0.79-0.93) | **<0.001** | 0.85 (0.79-0.91) | **<0.001** |

\*Continuous variables with outliers have been truncated at the 5<sup>th</sup> and 95<sup>th</sup> percentiles to reduce outlier influence.
<sup>†</sup>p-values are for a Wald test. p-values <0.05 are shown in **bold**.
Abbreviations: CIs – confidence intervals; IQR- interquartile range; ST- Multi-locus sequence type.

**Figure 3:**
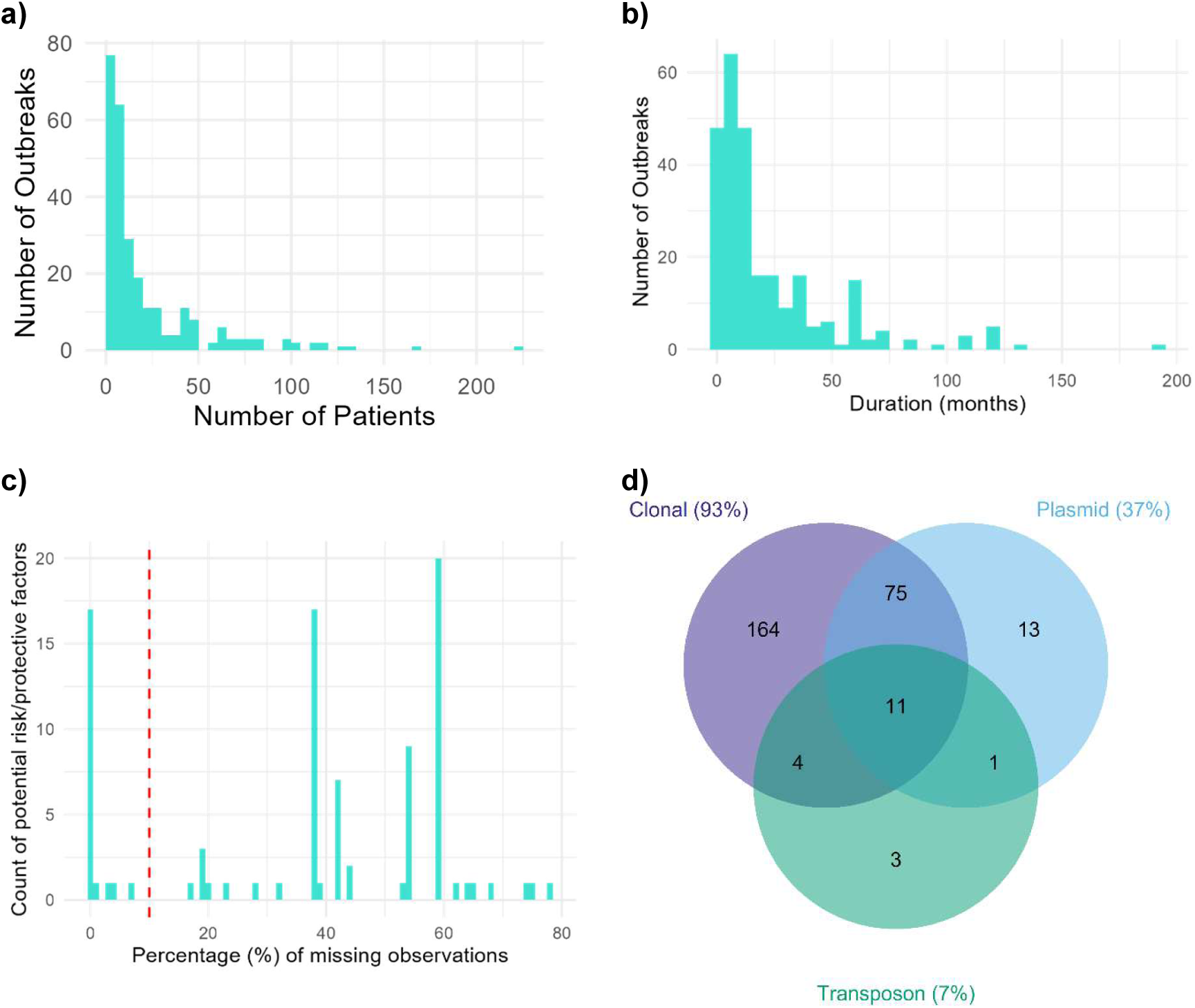
Distribution of outbreak outcomes, risk/protective factor missingness and main vectors of transmission. **a)** Distribution the primary outbreak outcome measure, outbreak size (total number of patients infected/ colonised per outbreak) among 271/272 outbreaks reporting outbreak size and **b)** distribution of the secondary outcome, outbreak duration, in months among the 263/272 outbreaks reporting duration. **c)** Histogram of the percentage of outbreaks with missing data (i.e. % of missing observations) for the 93 potential risk/protective factors considered. A vertical red dashed line is drawn at 10% missingness, which was the threshold used for inclusion of potential risk/protective factors as candidate variables in backwards selection for the final core models. Variables to the left of the red dashed line were included as candidate variables in backwards selection for the final core models. **d)** Venn diagram of the vectors of carbapenemase gene transmission (as reported by the author, or as implied by the involvement of different genomic contexts of the same carbapenemase allele, where not explicitly reported) involved in outbreaks, among 271 outbreaks with complete primary outcome data.

Most outbreaks were small, involving median 10 infected/colonised patients (IQR=5-27), lasting median 12 months (IQR=5-30 months; Table 2; Fig.3a/b). Outbreak follow-up period was not recorded, as this was sparsely reported during data extraction piloting. A greater proportion of affected patients were infected compared to colonised (median: 0.5 [IQR=0.22-1.0] vs 0.4 [0.0-0.79]), although infection/colonisation status was unavailable/unclear for 36% (2,198/6,175) of affected patients across included studies (Table 2). The median mortality rate was 30% (IQR=12-47%). Just over half the outbreaks (55% [82/149]) were ‘resolved’, with CPE infection/colonisation incidence returning to the author-defined ‘baseline’. ‘Resolved’ outbreaks were shorter (median 9 months [IQR=4-17] for 80/82 ‘resolved’ outbreaks with available duration data) than ‘unresolved’ outbreaks (median 24 months [IQR=10-47] for 66/67 ‘unresolved’ outbreaks with duration data; Wilcoxon ranksum *p<0.0001*).

Almost all outbreaks (93% [254/271]) reported carbapenemase gene transmission associated with clonal (i.e.lineage) spread, while 39.5% (107/271) reported MGE-mediated transmission (32% [88/271] plasmid-mediated only, 3% [7/271] transposon/IS/integron-mediated only, and 4% [12/271] involving both [+/- clonal transmission]; Table S4; Fig.3d). However, 66% (104/157) of studies providing an outbreak definition relied on clonal-based definitions, rather than gene- or plasmid-based definitions (Table S4). Additionally, 44% [110/249] of outbreaks used exclusively short-read sequencing (versus 53% [133/249] hybrid, 2% [6/249] long-read only, and 8% [22/271] with no sequencing modality reported), perhaps contributing to ascertainment bias. Epidemic plasmids (i.e. plasmids associated with carbapenemase gene dissemination either through clonal transmission or HGT) were described in 77% (208/271) of outbreaks (Table S5). Multi-replicon plasmids constituted 20% (36/184) of epidemic plasmids, and were collectively more abundant than any individual single-replicon plasmid (IncFIB and IncX both 12% [22/184], IncN 11% [21/184], IncL/M 8% [14/184], IncFIB 7% [12/184], and IncL 5% [10/184]). *K. pneumoniae* isolates were the most common, predominating in 63% (171/271) of outbreaks, particularly *K. pneumoniae* ST11, ST258, ST14, ST15 and ST307, while *E. cloacae* complex was the second most common, in 18% (48/271)(Table S5).

The final multivariable mixed effects linear regression model (n=256 outbreaks with complete data), fitted using backwards selection from all 21 variables with <10% missing data, retained six variables: involvement of clonal transmission, number of institutions, number of Enterobacterales STs, outbreak publication year, start year, and number of outbreaks (all *p<0.05*; Table 3; Figure 4a). Of the two collinear variables, number of Enterobacterales STs and number of species (ρ=0.86), the model including number of Enterobacterales STs had a lower AIC and was thus chosen as the core model.

**Figure 4:**
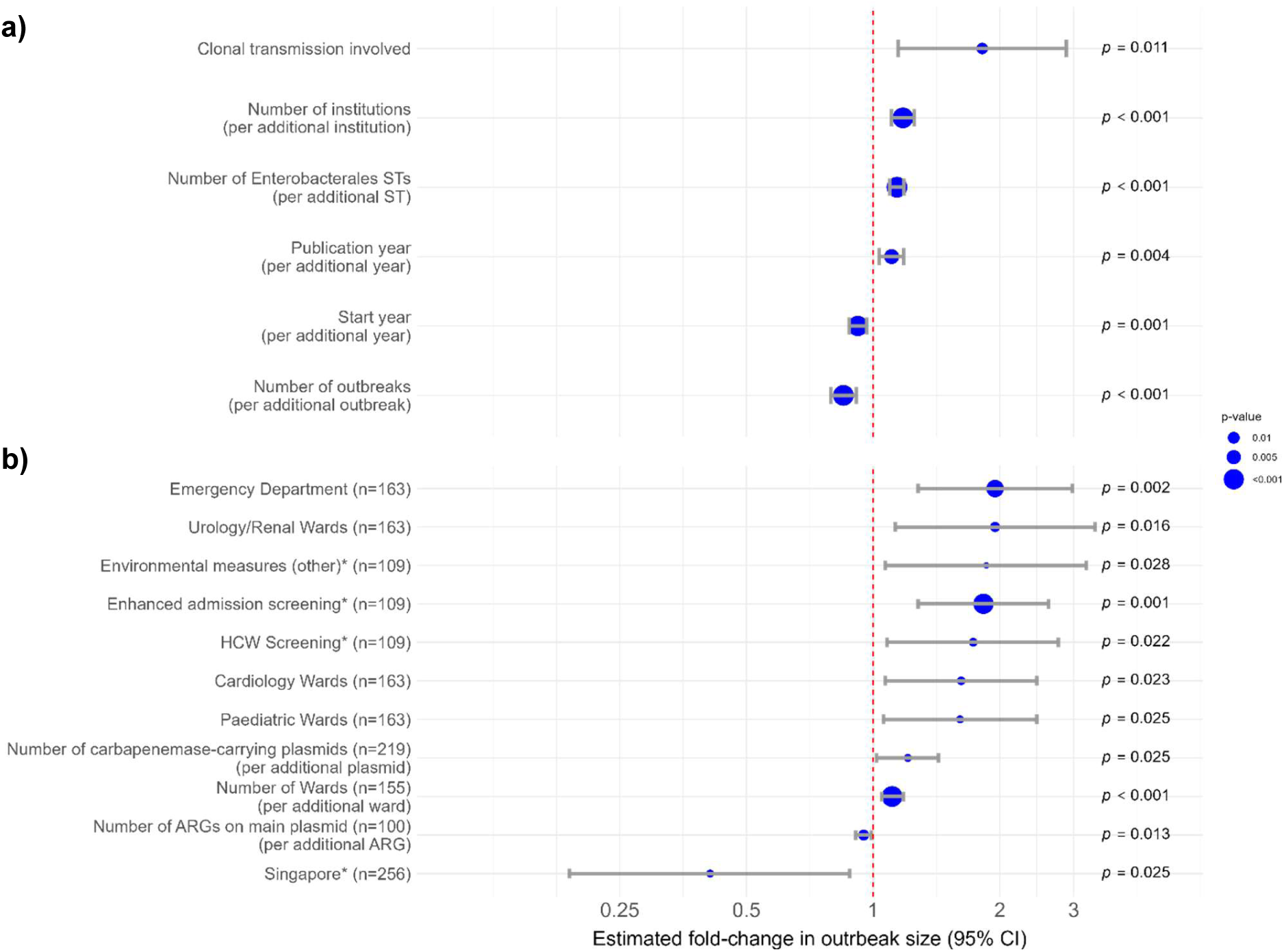
Fold-change in outbreak size associated with risk/protective factors a) included in the final core multivariable model (≥90% complete data) and b) not included in the final core multivariate model (either not retained in the core model through backwards selection or <90% complete data), but added back into the core model individually to obtain an adjusted estimate. In both **a)** and **b)**, each point estimated fold-change (circle) and 95% CIs (grey bars) are adjusted for all variables in the core model shown in **a)**. Only statistically significant risk/protective factors are shown in **b)**. The core model in **a)** is based on 256 observations (i.e. reported outbreaks) with complete data, while the number of observations (outbreaks) included in the adjusted models to obtain each estimated fold-change in **b)** varies and is shown for each variable in brackets after the variable name. *The reference category used for infection control measures was outbreaks not reporting that specific control measure, among outbreaks reporting any control measures. The reference category used for country of outbreak was China, as it had the largest number of reports. ^*^The reference category used for infection control measures was outbreaks not reporting that specific control measure, among outbreaks reporting any control measures. The reference category used for country of outbreak was China, as it had the largest number of reports.

The involvement of clonal transmission was associated with larger outbreaks (relative size: 1.82 [95% CI: 1.15-2.88]; *p=0.011*). The involvement of more institutions (1.18 per institution [1.11-1.25]; *p<0.001*), more Enterobacterales STs (1.14 per ST [1.10-1.19]; *p<0.001*), and later publication year (1.11 per year [1.03-1.18]; *p=0.004*) were also associated with larger outbreaks. Conversely, earlier start year (0.92 per year [0.88-0.97]; *p=0.001*) and reporting more outbreaks in the study (0.85 per outbreak [0.79-0.91]; *p<0.001*) were associated with smaller outbreaks (Table 3; Figure 4a).

When each other variable with ≥12 datapoints (<90% completeness or not retained in the core model) was individually added into the core model, 11 variables had significant effects after adjusting for the six core model variables (Tables S3-6; Fig.4b). The involvement of emergency departments, urology/renal wards, cardiology and paediatric wards, and reporting ‘other’ environmental controls (environmental measures other than screening/cleaning/replacement or ward closure; Table S6), enhanced patient admission screening, healthcare worker (HCW) screening, and more carbapenemase-associated plasmids were associated with larger outbreaks, while having more ARGs on the outbreak plasmid, and occurring in Singapore (versus China) was associated with smaller outbreaks (Tables S3-6; Fig.4b). Importantly, due to missing data, some of these models were fitted on smaller data subsets (100-256 outbreaks; Fig.4b).

A sensitivity analysis restricted regression analyses to outbreaks affecting ≥10 patients (‘10-or-more-patients’ outbreak model; see Methods). Seven variables were retained in this core model (number of institutions, number of Enterobacterales STs, number of outbreaks reported in the study, as in the above model, plus main ST, publication type, involvement of transposon/IS-mediated transmission, number of carbapenemase alleles involved; n=133 complete outbreak records; Table S7; Fig.S2a). As in the all-outbreaks core model, in the 10-or-more-patients outbreak core model, more institutions and more Enterobacterales STs were associated with larger outbreaks (relative size: 1.10 per institution [95% CI: 1.04-1.16]; *p=0.001* and 1.04 per ST [1.01-1.08]; *p=0.011*), while number of outbreaks reported was associated with smaller outbreaks (0.93 per outbreak [0.87-0.99]; *p=0.031*). Publication type (abstracts versus peer-reviewed articles) (1.97 [1.22-3.17]; *p=0.06*), transposon/IS mediated transmission (1.53 [1.03-2.26]; *p=0.035*), and the involvement of *E. coli* STs (2.13 [1.28-3.56]; *p=0.004*), *K. pneumoniae* ST11 (1.66 [1.13-2.42]; *p=0.010*) and *K. pneumoniae* ST14 (1.82 [1.08-3.07]; *p=0.024*; versus ‘other/minor’ *K. pneumoniae* STs) were associated with larger outbreaks only in the 10-or-more-patients model (Table S7; Fig.S2a).

When other variables were added to the 10-or-more-patients model individually, 11 variables were significant (Fig.S2b). The reporting of HCW screening, environmental screening, cardiology and renal/urology wards, and government funding was associated with larger outbreaks, while more ARGs and small MGEs on the main plasmid, larger main plasmid size, clonal outbreak definition, and background screening were associated with smaller outbreaks (Table S7-10; Fig.S2b).

### Study reporting quality assessment

We targeted 100 studies at random for reporting quality assessment of which two were recognised as ineligible on data extraction and excluded. Reporting quality was therefore assessed for a randomly selected 98/179 (55%) of studies (51% [140/272] of outbreaks; median outbreak size 10 patients [IQR=5-26]; Wilcoxon ranksum versus non-assessed outbreaks *p=0.604*). Studies reported adequately on median 11/19 relevant ORION items (IQR=8-13; range=1-18)(Fig.5). Reporting of the Title/abstract, Introduction, and Discussion were good (≥50% studies reporting adequately [Fig. 5; Table S12]).

**Figure 5:**
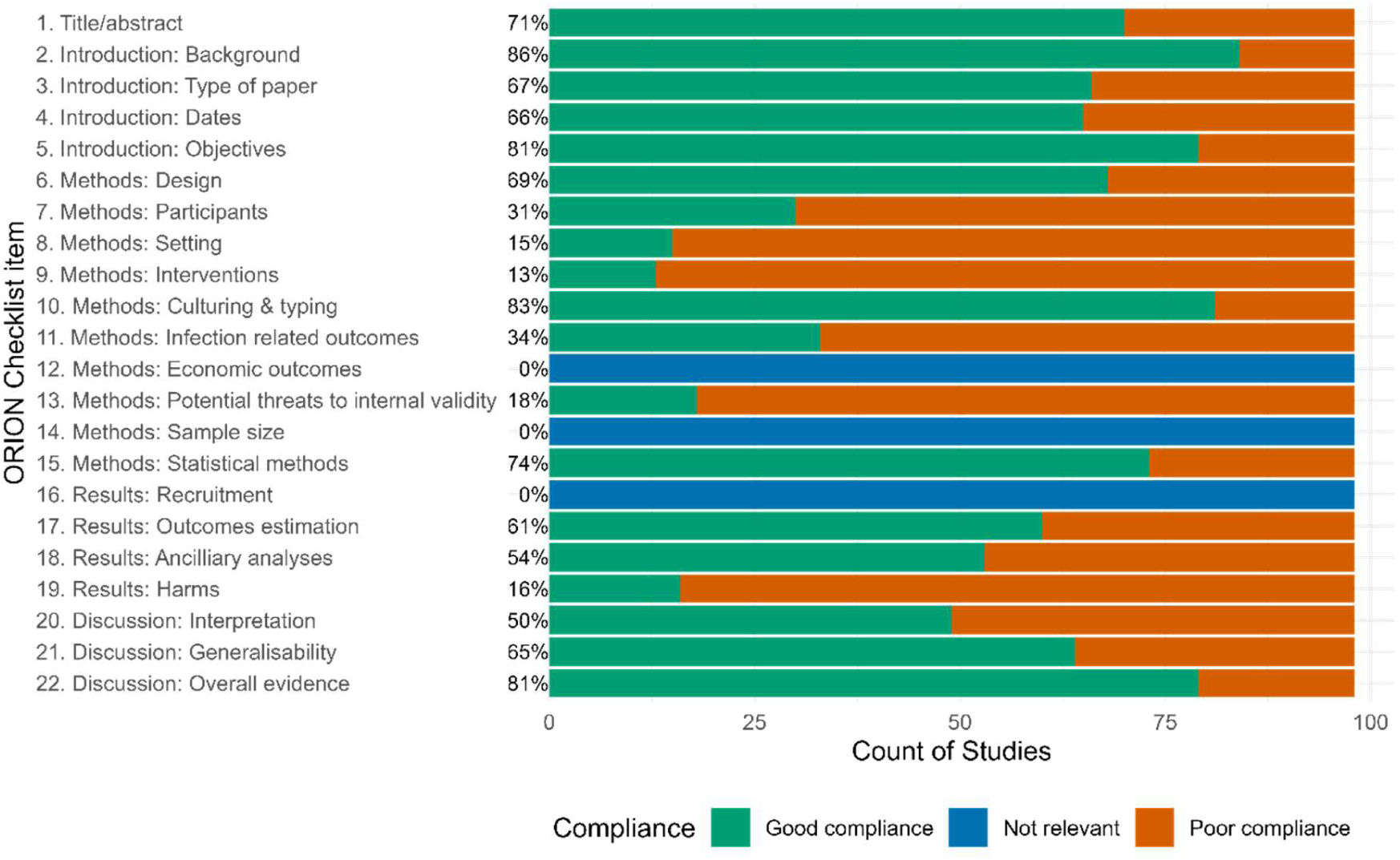
Compliance with the Outbreak Reports and Intervention Studies of Nosocomial Infection (ORION) checklist(26, 27) items for 98 studies for which quality of reporting was assessed.

Reporting of methods and results was variable, with better reporting of study design, culture/typing methods, statistical methods, outcome estimation, and ancillary analyses versus poor reporting on participants, settings, interventions, infection-related outcomes, potential threats to internal validity, and harms (Fig.5; Table S12).

## Discussion

Our review of 272 reported healthcare-associated CPE outbreaks over the last 20 years highlights their diverse dynamics, and that these are a global challenge. The most common ‘big five’ carbapenemase genes involved in outbreaks were *bla_KPC_* (31%), *bla*_NDM_ (29%), and *bla*_OXA-48-like_ (19%), in specific *K. pneumoniae* STs (ST11/14/15/258/307), as well as *E. cloacae*, and *E. coli*, although with regional variation (Fig.2). Our objective was to use reported data to identify factors predicting increased outbreak potential to support frameworks for managing healthcare-associated CPEs(31–33) by enabling early targeting of IPC resource at institutional and public health levels. A major challenge however was the variability in definitions used across reported studies (e.g. outbreak definitions), likely reporting bias (e.g. under-ascertainment of plasmid/MGE-associated or over-estimation of mortality) and likely reverse causality associated with some findings (e.g. reporting HCW/patient screening being associated with larger outbreaks).

The involvement of more institutions and more Enterobacterales STs was associated with larger outbreaks, supporting the intuitive fact that evidence of inter-institutional and inter-lineage horizontal gene transfer (HGT) represent key red flags for dissemination potential - although these may also reflect different sampling strategies driven by the perception of risk posed. The ‘protective’ effect of reporting more outbreaks in a study may reflect reporting differences, with larger outbreaks more likely to have entire studies dedicated to their reporting.

In the ‘all-outbreaks’ model, clonal transmission and later publication year were additional risk factors for larger outbreaks, and later start year was protective (Fig.4; Table 3). Earlier start year and later publication year may be indicators for outbreak duration, allowing more time for widespread transmission. The association with later publication year may also suggest that larger outbreaks are increasingly occurring or being reported. In the ’10-or-more-patients’ model, reporting in an abstract compared to a journal article, the involvement of certain species/STs (*E. coli* STs, ST14 and ST11 *K. pneumoniae* compared to other/minor *Klebsiella* STs) and transposon/IS-mediated transmission (Fig.S2; Table S7) were additional risk factors. These estimates had high uncertainty, with wide 95% CIs and/or lower/upper CIs close to the null, cautioning about inference regarding these factors from this dataset. However, considering the microbiological/genomic associations, *E. coli* carriage is near-ubiquitous in humans, and therefore carbapenemases acquired by this species may be a particular risk for dissemination. Additionally, *K. pneumoniae* lineages ST11 and ST14 are recognised as global ‘problem’ clones with respect to broader (including non-carbapenem) drug resistance and dissemination capacity(34). Transposons and ISs may enable the rapid transfer of carbapenemase genes amongst and rearrangement in different genetic backgrounds, thereby facilitating adaptation for dissemination and persistence. Nonetheless, the significance of several study-level variables, such as publication type, highlights the potentially important confounding effect of outbreak reporting, and the reporting context should therefore be considered when interpreting outbreak reports.

Four additional variables with higher data missingness were associated with larger outbreaks in both multivariable models: the involvement of urology/renal and cardiology wards, reporting HCW screening, and fewer ARGs on the main outbreak plasmids. Urology/renal wards, cardiology wards, and HCW screening had moderate effect sizes, each resulting in 50-95% larger outbreaks when present, while each extra ARG present on the outbreak plasmid was associated with 4-5% smaller outbreaks. The larger outbreak size associated with urology/renal wards echoes findings from observational studies where renal patients had higher rates of colonisation with CPEs(35–37), and are also a ‘high-risk’ group in national(31) and international(32) CPE guidelines. Cardiology patients may also have many known risk factors for CPE colonisation/infection, including co-morbidity, recent invasive/surgical procedure and indwelling medical devices(31, 38–40). Notwithstanding the risk of confirmation bias (i.e. where known risk factors are more likely to be investigated/reported), cases in these patient groups may represent an early signal of outbreak potential. The association between reporting HCW screening and larger outbreaks likely reflects reverse causality, where larger outbreaks result in expanded control measures.

The protective effect of more ARGs on the main outbreak plasmid may be related to an increased fitness cost of carrying more ARG ‘cargo’, both for clonal transmission(41–43), and HGT(44). However, many AMR gene/plasmid combinations may have no fitness cost due to compensatory mutations/co-evolution, or fine-tuning expression(45). Reporting bias is thus also plausible, whereby small outbreaks of highly-resistant lineages are more likely to be reported than less resistant strains, or whereby more bioinformatic expertise may be available to characterise plasmids in settings that are better-resourced to control transmission. Importantly, high missingness raises the risk of bias due to non-randomly missing data.

Eight variables were only significant in the ‘all-outbreaks’ multivariable model (emergency department/paediatric ward involvement, ‘other’ environmental controls, enhanced patient admissions screening, more carbapenemase-carrying plasmids, and more wards were risk factors, while occurring in Singapore versus China was protective). Seven variables were significant only when adjusted using the ‘10-or-more-patients’ outbreak model (*bla*_OXA-48-like_ as the main gene family and environmental screening were risk factors, while reporting more small MGEs on the main outbreak plasmid, receiving government funding, clonal outbreak definitions, and background screening were protective). However, these adjusted estimates were not robust across both multivariable models, were based on smaller sample sizes due to missing data, and had high uncertainty.

To our knowledge, this is the first quantitative synthesis of CPE outbreak risk factors across several domains (epidemiological, microbiological/genomic, and infection control measures) using almost 300 outbreak reports. The absence of geographical/temporal restrictions offers a global perspective and enhances statistical power.

The study may be affected by selection bias from using WGS as an inclusion criterion, possibly excluding outbreaks in lower-resource settings. However, this was necessary to consider relevant genetic aspects. This may be reflected in the predominance of studies from the global North and China, limiting generalisability. Although attempts were made to capture diverse potential confounders, this dataset was not sufficiently powered to detect interactions or allow subgroup analyses. To maintain statistical power, under a third of recorded covariates were considered in the final backwards selection models, risking residual confounding. Furthermore, extraction for the full study set was done by a single reviewer, thus potentially reflecting their subjectivity in interpreting/categorising variables. Outbreak reporting of ORION checklist items relating to participants/settings, interventions, infection-related outcomes, internal validity was inconsistent (Fig.5; Table S12). This emphasises the limitations of inferences from available data, especially relating to epidemiological/patient-level metadata, despite significant publication efforts to date.

In conclusion, this systematic review highlights the diversity of healthcare-associated CPE outbreaks. At least 40% of outbreaks involved MGE-mediated transmission, and outbreaks tended to be larger when more institutions or more Enterobacterales STs were involved. Nevertheless, incomplete and heterogenous reporting pose challenges to integrated risk quantification of epidemiological, microbiological, and genomic factors using publicly available literature. Systematic sampling, sequencing and evaluation of associated microbiological and patient-level metadata to characterise background and outbreak epidemiology will be essential to develop healthcare-associated CPE dissemination risk frameworks.

## Data Availability

The cleaned extraction dataset and analysis scripts are available online (https://figshare.com/articles/dataset/Extraction_data_and_R_analysis_script_for_CPE_systematic_review/30148717).

## Ethical considerations

This study analysed publicly available reports and no additional ethical review was sought.

## Transparency Declaration Conflict of Interest

All authors have no conflicts of interest to declare.

## Acknowledgements

An earlier version of this work was presented as an academic poster at the Congress of the European Society of Clinical Microbiology and Infectious Diseases (ESCMID Global) 2025.

## Access to Data

The extracted dataset and R analysis scripts has been uploaded to FigShare (https://figshare.com/articles/dataset/Extraction_data_and_R_analysis_script_for_CPE_systematic_review/30148717). The full search strategy can be found in the Supplementary materials. The review has been registered on PROSPERO (<u>CRD42024505048</u>).

## Contributions

WM, SH, ASW, AL, JR, NS, and SL were involved in conceptualisation, funding acquisition, project administration, provision or resources and supervision. DN, AB, CBS, JY were involved in screening, extraction, data curation and validation under the supervision of NS and SL. DN, SL and NS were involved with methodological development, formal analysis, investigation, visualisation and writing the original draft and reviewing/editing. All authors approved the final draft.

