## Supplementary material for "Epidemiological, microbiological, and genomic risk factors for healthcare-associated Carbapenemase producing Enterobacterales (CPE) outbreaks: A systematic review": Table S1; Table S2; Table S3; Table S4; Table S5; Table S6; Table S7; Table S8; Table S9; Table S10; Table S11; Table S12; Fig.S1

**Table S1: Search records for systematic review including source and date or latest search strategy and number of records retrieved.**

| Source Type | Source | Date | Search Strategy | Records Retrieved |
|---|---|---|---|---|
| Database | MEDLINE | 21/12/23 | Medline (Ovid MEDLINE® Epub Ahead of Print, In-Process & Other Non-Indexed Citations, Ovid MEDLINE® Daily and Ovid MEDLINE®) 1946 to present<br>1 Carbapenem-Resistant Enterobacteriaceae/ 1523<br>2 ((carbapenem* adj3 resist*) or (carbapenem* adj3 non-suscept*) or carbapenemase*).ti,ab,kw. 16855<br>3 ("NDM" or "KPC" or "VIM" or "IMP" or MET?1 or "OXA-48" or "OXA48" or "OXA-48-like" or OXA-162 or OXA-181 or OXA-199 or OXA-204 or OXA-232 or OXA-244 or OXA-245 or OXA-370 or OXA-517 or OXA-1038).ti,ab,kw. 20002<br>4 exp Disease Outbreaks/ or (Disease Transmission, Infectious/ or Infectious Disease Transmission, Patient-to-Professional/ or Infectious Disease Transmission, Professional-to-Patient/) or Cross Infection/ 295736<br>5 (outbreak* or epidemic* or (case* adj3 cluster*) or (case* adj3 series) or (infect* adj3 cluster*) or (infect* adj3 series) or epidem* cluster* or (prolong* adj3 transmiss*)).ti,ab,kw. 394711<br>6 Plasmids/ or F Factor/ or R Factors/ or (Conjugation, Genetic/ or Gene Transfer, Horizontal/) 125371<br>7 (plasmid* or F-factor or R-factor or integrative conjugative element or horizontal gene* transfer or horizontal gene* exchange or mobil* gen* element* or insertion sequence or integron or transposon or replicon or gene cassette or cassette array).ti,ab,kw. 182445<br>8 1 or 2 16931<br>9 1 or 2 or 3 30969<br>10 3 and 8 5964<br>11 4 or 5 593090<br>12 6 or 7 238775<br>13 9 and 11 3704<br>14 10 and 11 1213<br>15 9 and 12 4885<br>16 10 and 12 2207<br>17 9 and 11 and 12 973 | 973 |
| Database | Embase | 21/12/23 | Embase 1974 to present<br>1 exp carbapenem resistance/ 2239<br>2 ((carbapenem* adj3 resist*) or (carbapenem* adj3 non-suscept*) or carbapenemase*).ti,ab,kw. 22120<br>3 ("NDM" or "KPC" or "VIM" or "IMP" or MET?1 or "OXA-48" or "OXA48" or "OXA-48-like" or OXA-162 or OXA-181 or OXA-199 or OXA-204 or OXA-232 or OXA-244 or OXA-245 or OXA-370 or OXA-517 or OXA-1038).ti,ab,kw. 26823<br>4 exp epidemic/ 132962<br>5 disease transmission/ or horizontal disease transmission/ or indirect contact transmission/ or nosocomial transmission/ or pathogen transmission/ 116513<br>6 cross infection/ 20824<br>7 patient-to-patient transmission/ or patient-to-professional transmission/ 84<br>8 professional-to-patient transmission/ 14<br>9 (outbreak* or epidemic* or (case* adj3 cluster*) or (case* adj3 series) or (infect* adj3 cluster*) or (infect* adj3 series) or epidem* cluster* or (prolong* adj3 transmiss*)).ti,ab,kw. 470432<br>10 exp plasmid/ 145990<br>11 bacterium conjugation/ 4863<br>12 horizontal gene transfer/ 9676<br>13 exp mobile genetic element/ 191431<br>14 (plasmid* or F factor or R factor or integrative conjugative element or horizontal gene* transfer or horizontal gene* exchange or mobil* gen* element* or insertion sequence or integron or transposon or replicon or gene cassette or cassette array).ti,ab,kw. 209790<br>15 1 or 2 or 3 41929<br>16 4 or 5 or 6 or 7 or 8 or 9 618644<br>17 10 or 11 or 12 or 14 265205<br>18 10 or 11 or 12 or 13 or 14 292020<br>19 15 and 16 and 17 931<br>20 15 and 16 and 18 964 | 964 |

| Source Type | Source | Date | Search Strategy | Records Retrieved |
|---|---|---|---|---|
| Database | Scopus (through Science direct) | 22/12/23 | ( TITLE-ABS-KEY ( ( carbapenem* W/3 resist* ) OR carbapenem-resist* OR ( carbapenem* W/3 non-suscept* ) OR carbapenemase* ) OR ALL ( "NDM" OR "KPC" OR "VIM" OR "IMP" OR met-1 OR "OXA-48" OR "OXA48" OR oxa-162 OR oxa-181 OR oxa-199 OR oxa-204 OR oxa-232 OR oxa-244 OR oxa-245 OR oxa-370 OR oxa-517 OR oxa-1038 ) AND TITLE-ABS-KEY ( outbreak OR epidemic OR ( case* W/3 cluster* ) OR ( case* W/3 series ) OR ( infect* W/3 cluster* ) OR ( infect* W/3 series ) OR ( epidem* AND cluster* ) OR ( prolong* W/3 transmiss* ) ) AND TITLE-ABS-KEY ( plasmid* OR f-factor OR r-factor OR (integrative conjugative element) OR (horizontal gene transfer) OR (horizontal transfer of genes) OR (horizontal gene exchange) OR (horizontal genetic exchange) OR (mobile genetic element) OR (insertion sequence) OR integron OR transposon OR replicon OR (gene cassette) OR (cassette array) ) ) | 1304 |
| Database | Web of Science (through Clarivate) | 21/12/23 | Search #8<br>**TS=("NDM" or "KPC" or "VIM" or "IMP" or MET$1 or OXA$48 or OXA-162 or OXA-181 or OXA-199 or OXA-204 or OXA-232 or OXA-244 or OXA-245 or OXA-370 or OXA-517 or OXA-1038)** and **Preprint Citation Index** (Exclude – Database)<br>Search #9<br>**TS=(outbreak* or epidemic* or (case* NEAR/3 cluster*) or (case* NEAR/3 series) or (infect* NEAR/3 cluster*) or (infect* NEAR/3 series))** and **Preprint Citation Index** (Exclude – Database)<br>Search #10<br>**TS=(plasmid* or F-factor or R-factor or (horizontal gene* transfer) or (horizontal gene* exchange) or (mobil* gen* element*) or integron or transposon)** and **Preprint Citation Index** (Exclude – Database)<br><br>#8 AND #9 AND #10<br>Notes:Data sets and retracted papers excluded<br>Searched all in 'Topic' field tag, which searches abstract, title, and indexing by individual databases belonging to Web of Science | 993 |
| Database | Global Health (through OVID) | 21/11/23 | Global Health <1973 to 2023 Week 51><br>1 beta-lactamase/ 11181<br>2 antibiotic resistance/ 9279<br>3 ((carbapenem* adj3 resist*) or (carbapenem* adj3 non-suscept*) or carbapenemase*).ti,ab. 11804<br>4 ("NDM" or "KPC" or "VIM" or "IMP" or MET?1 or "OXA-48" or "OXA48" or "OXA-48-like" or OXA-162 or OXA-181 or OXA-199 or OXA-204 or OXA-232 or OXA-244 or OXA-245 or OXA-370 or OXA-517 or OXA-1038).ti,ab. 6562<br>5 outbreaks/ 56625<br>6 exp epidemics/ 100228<br>7 disease transmission/ or horizontal transmission/ or infection/ or occupational transmission/ 97687<br>8 cross infection/ 541<br>9 (outbreak* or epidemic* or (case* adj3 cluster*) or (case* adj3 series) or (infect* adj3 cluster*) or (infect* adj3 series) or epidem* cluster* or (prolong* adj3 transmiss*)).ti,ab. 158588<br>10 exp plasmids/ 10003<br>11 exp transposable elements/ 4486<br>12 gene transfer/ 2392<br>13 (plasmid* or F factor or R factor or integrative conjugative element or horizontal gene* transfer or horizontal gene* exchange or mobil* gen* element* or insertion sequence or integron or transposon or replicon or gene cassette or cassette array).ti,ab. 31503<br>14 3 or 4 14391<br>15 1 or 3 or 4 21773<br>16 1 or 2 or 3 or 4 29543<br>17 5 or 6 or 7 or 8 or 9 293912<br>18 10 or 11 or 12 or 13 34508<br>19 14 and 17 and 18 625<br>20 15 and 17 and 18 836 | 836 |

| Source Type | Source | Date | Search Strategy | Records Retrieved |
|---|---|---|---|---|
| Database | BioRxiv/ MedRxiv | 23/12/23 | Due to limited capacity to combine Boolean operators (AND and OR) on search interface, each pairwise combination of the following two lists was searched in the title and abstract fields:<br>(OXA, KPC, NDM, VIM, IMP, carbapenemase, carbapenem resistant, carbapenem resistance, carbapenem-resistant, carbapenem-resistance, carbapenem npn-susceptible, carbapenem-non-susceptible)<br>AND<br>(outbreak, epidemic, cluster, spread, series, prolonged transmission) | 135 |
| Outbreak registry/ surveillance | ECDC Reports | 17/01/24 | carbapenemase<br><br>Notes: news, corporate documents and letters removed | 139 |
| | CDC MMWR | 24/12/23 | carbapenem | 14 |
| | WWDNO | 20/12/23 | "carbapenem" OR "carbapenemase" | 50 (+55 publications) |
| Conferences | IDSA IDWeek | 20/01/24 | carbapenemase AND outbreak<br>Notes:3 images removed | 178 |
| | ASM Microbe | 18/01/24 | carbapenemase<br><br>Notes:2019-2023<br>Abstracts only | 161 |
| | ECCMID | 18/01/24 | "carbapenemase" AND "outbreak"<br><br>Notes: 2012-2023 | 106 |

**Abbreviations**: ECDC- European Centre of Disease Control and Prevention; CDC- US Centre of Disease Control and Prevention; MMWR- Morbidity and mortality weekly reports; WWDNO- Worldwide Database for Nosocomial Outbreaks; IDSA IDWeek- Infectious Disease Society of America Infectious Disease Week; ASM- American Society of Microbiology; ECCMID- European Congress of Clinical Microbiology and Infectious Diseases

**Table S2: Summary of study-level characteristics of 179 de-duplicated studies included in the systematic review.**

| Variable / Categories | Missing (%) | Number of studies (%) with data available | Median (IQR) or Number (%) of studies | Range |
|---|---|---|---|---|
| **Publication year** | 0 (0.0%) | 179 (100%) | 2020 (2018-2022) | 2013-2024 |
| **Publication type** | 0 (0.0%) | 179 (100%) | | |
| Peer-reviewed journal article | | | 157 (87.7%) | - |
| Abstract | | | 19 (10.6%) | - |
| Preprint | | | 3 (1.7%) | - |
| **Study design (main)** | 0 (0.0%) | 179 (100%) | | |
| Outbreak report | | | 115 (64.2%) | - |
| Retrospective descriptive | | | 57 (31.8%) | - |
| Prospective descriptive | | | 4 (2.2%) | - |
| Case-control | | | 2 (1.1%) | - |
| Cross-sectional | | | 1 (0.6%) | - |
| **Funding†** | 37 (20.7%) | 142 (79.3%) | | |
| Government | | | 119 (83.8%) | - |
| Academic | | | 36 (25.4%) | - |
| Other | | | 28 (19.7%) | - |
| **Conflict of interest** | 33 (18.4%) | 146 (81.6%) | | |
| Absent | | | 127 (87.0%) | - |
| Present | | | 19 (13.0%) | - |
| **Number of outbreaks reported in study** | 0 (0.0%) | 179 (100%) | 1 (1-1) | 1-9 |

* Percentages are calculated using the denominator of the number of studies with data available (non-missing) for that variable.
†Funding sources were recorded as a non-mutually exclusive categorical variable, allowing for a sum >100%.

**Table S3: Study-level risk/protective factors for outbreak size with descriptive summary, univariable and multivariable associations with outbreak size (total number of patients infected/colonised per outbreak).** Descriptive summaries are provided considering the dataset of 271 outbreaks with complete outcome data (out of 272 included in this review). Univariable associations are calculated from a dataset of outbreaks with complete data for each variable (shown in the 3rd column from the left). Multivariable associations are calculated from a dataset of outbreaks with complete data on that variable as well as all of the variables included in the final model (i.e. number of outbreaks reported in study, publication year, number of institutions, start year, number of STs involved, clonal transmission involved). Estimates and 95% confidence intervals (CIs) are reported on the original (non-$log_{10}$) scale, interpreted as relative changes in outbreak size (fold-change in size).

| Variable / Category | Number (%) missing | Number of (%) outbreaks with data available | Median (IQR) or Number (%) | Range | Median (IQR) number of patients affected | Univariable Fold-change (95% CIs) | p-value† | Multi-variable N | Multivariable‡ Fold-change (95% CIs) | p-value† |
|---|---|---|---|---|---|---|---|---|---|---|
| **Publication year** | 0 (0%) | 271 (100%) | 2021 (2019-2022) | 2016-2023 | - | 1.04 (0.97-1.11) | 0.248 | 256 | 1.11 (1.03-1.18) | 0.004 |
| **Publication type** | | | | | | | | | | |
| **Journal article** | 0 (0%) | 271 (100%) | 237 (87.5%) | - | 10 (5-26) | *ref* | 0.397 | - | *ref* | 0.699 |
| **Abstract** | - | - | 31 (11.4%) | - | 7 (4-28) | 0.76 (0.46-1.27) | 0.290 | 256 | 1.18 (0.78-1.79) | 0.431 |
| **Preprint** | - | - | 3 (1.1%) | - | 15 (13-46) | 1.70 (0.47-6.22) | 0.420 | 256 | 1.21 (0.42-3.50) | 0.728 |
| **Study design** | | | | | | | | | | |
| **Outbreak report** | 0 (0%) | 271 (100%) | 137 (50.6%) | - | 10 (5-25) | *ref* | 0.301 | - | *ref* | 0.612 |
| **Case-control** | - | - | 6 (2.2%) | - | 5 (4-10) | 0.50 (0.14-1.78) | 0.282 | 256 | 0.69 (0.26-1.86) | 0.460 |
| **Cross-sectional** | - | - | 1 (0.4%) | - | 115 (115-115) | 8.56 (0.92-79.72) | 0.059 | 256 | 2.90 (0.46-18.46) | 0.258 |
| **Prospective descriptive** | - | - | 6 (2.2%) | - | 14 (6-31) | 1.17 (0.42-3.27) | 0.768 | 256 | 1.41 (0.64-3.13) | 0.393 |
| **Retrospective descriptive** | - | - | 121 (44.6%) | - | 10 (4-28) | 1.00 (0.72-1.39) | 1 | 256 | 0.97 (0.72-1.31) | 0.842 |
| **Funding** | | | | | | | | | | |
| **Government funding*** | 51 (19%) | 220 (81%) | 194 (88.2%) | - | 10 (5-27) | 0.79 (0.48-1.28) | 0.336 | 207 | 0.87 (0.59-1.27) | 0.458 |
| **Academic funding*** | - | - | 45 (20.5%) | - | 14 (7-40) | 1.33 (0.88-2.02) | 0.170 | 207 | 1.26 (0.92-1.74) | 0.148 |
| **Other funding*** | - | - | 40 (18.2%) | - | 9 (6-33) | 1.15 (0.74-1.79) | 0.541 | 207 | 1.01 (0.72-1.42) | 0.943 |
| **Conflict of interest** | | | | | | | | | | |
| **Declared present*** | 54 (20%) | 217 (80%) | 28 (12.9%) | - | 13 (7-36) | 1.10 (0.66-1.82) | 0.708 | 203 | 1.01 (0.70-1.46) | 0.960 |
| **Number of outbreaks reported in publication** | 0 (0%) | 271 (100%) | 1 (1-4) | 1-9 | - | 0.86 (0.79-0.93) | **<0.001** | 256 | 0.85 (0.79-0.91) | **<0.0001** |

* Compared to the absence of reporting that type of funding or conflict of interest, for those outbreaks where funding and conflict of interest was reported, respectively.

†p-values are for a Wald test. p-values appearing in the reference ('ref') category row represent overall Wald-test p-values for that category, obtained using Satterthwaite's method for a type III ANOVA for mixed effects models. p-values <0.05 are shown in **bold**.

‡ Multivariable associations are adjusted for all 6 variables in the final core model: number outbreaks reported in study, publication year, number of institutions, start year, number of STs involved, clonal transmission involved. Abbreviations: IQR- interquartile range; CIs – confidence intervals.

**Table S4: Epidemiological risk/protective factors for outbreak size with descriptive summary, univariable and multivariable associations with outbreak size (total number of patients infected/colonised per outbreak).** Descriptive summaries are provided considering the dataset of 271 outbreaks with complete outcome data (out of 272 included in this review). Univariable associations are calculated from a dataset of outbreaks with complete data for each variable (shown in the 3rd column from the left). Multivariable associations are calculated from a dataset of outbreaks with complete data on that variable as well as all of the variables included in the final model. Estimates and 95% confidence intervals (CIs) are reported on the original (non-$log_{10}$) scale, interpreted as relative changes in outbreak size (fold-change in size).

| **Variable** / **Category** | **Number (%) missing** | **Number (%) outbreaks with data available** | **Median (IQR) or Number (%)** | **Range** | **Median (IQR) number patients affected** | **Univariable Fold-change (95% CIs)** | **p-value†** | **Multi-variable N** | **Multivariable‡ Fold-change (95% CIs)** | **p-value†** |
|---|---|---|---|---|---|---|---|---|---|---|
| **Start year** | 3 (1.1%) | 268 (98.9%) | 2016 (2014-2018) | 2010-2021 | - | 0.94 (0.9-0.98) | 0.007 | 256 | 0.92 (0.88-0.96) | **0.004** |
| **Continent** | | | | | | | | | | |
| **Europe** | 0 (0%) | 271 (100%) | 127 (46.9%) | - | 11 (6-36) | *ref* | 0.331 | - | *ref* | 0.660 |
| **Africa** | - | - | 7 (2.6%) | - | 19 (7-34) | 1.06 (0.43-2.63) | 0.901 | 256 | 0.80 (0.39-1.63) | 0.535 |
| **Asia** | - | - | 64 (23.6%) | - | 8 (4-19) | 0.71 (0.48-1.05) | 0.083 | 256 | 0.86 (0.62-1.18) | 0.342 |
| **Australasia** | - | - | 18 (6.6%) | - | 11 (7-46) | 1.05 (0.55-1.98) | 0.891 | 256 | 0.92 (0.55-1.55) | 0.751 |
| **North America** | - | - | 37 (13.7%) | - | 8 (4-15) | 0.64 (0.40-1.03) | 0.066 | 256 | 0.72 (0.49-1.05) | 0.087 |
| **South America** | - | - | 18 (6.6%) | - | 16 (6-32) | 0.93 (0.49-1.77) | 0.829 | 256 | 0.87 (0.53-1.42) | 0.567 |
| **Country** | | | | | | | | | | |
| **China** | 0 (0%) | 271 (100%) | 38 (14%) | - | 8 (5-23) | *ref* | 0.012 | - | *ref* | 0.081 |
| **Australia** | - | - | 18 (6.6%) | - | 11 (7-46) | 1.41 (0.72-2.74) | 0.316 | 256 | 0.94 (0.54-1.62) | 0.817 |
| **Canada** | - | - | 12 (4.4%) | - | 4 (2-5) | 0.46 (0.19-1.14) | 0.093 | 256 | 0.52 (0.25-1.10) | 0.085 |
| **Denmark** | - | - | 19 (7%) | - | 4 (2-10) | 0.47 (0.19-1.13) | 0.092 | 256 | 1.08 (0.48-2.42) | 0.844 |
| **Italy** | - | - | 11 (4.1%) | - | 23 (16-48) | 1.87 (0.88-3.98) | 0.102 | 256 | 1.69 (0.93-3.07) | 0.085 |
| **Other** | - | - | 75 (27.7%) | - | 14 (6-30) | 1.20 (0.77-1.87) | 0.425 | 256 | 0.87 (0.60-1.25) | 0.449 |
| **Poland** | - | - | 19 (7%) | - | 13 (7-57) | 2.26 (0.98-5.24) | 0.056 | 256 | 0.90 (0.41-1.98) | 0.786 |
| **Singapore** | - | - | 11 (4.1%) | - | 2 (2-3) | 0.37 (0.14-1.03) | 0.056 | 256 | 0.41 (0.19-0.88) | **0.025** |
| **Spain** | - | - | 20 (7.4%) | - | 14 (9-42) | 1.74 (0.88-3.44) | 0.108 | 256 | 1.37 (0.80-2.36) | 0.251 |
| **UK** | - | - | 23 (8.5%) | - | 7 (4-26) | 1.27 (0.64-2.53) | 0.486 | 256 | 0.85 (0.49-1.47) | 0.558 |
| **USA** | - | - | 25 (9.2%) | - | 12 (7-17) | 1.02 (0.58-1.79) | 0.953 | 256 | 0.81 (0.51-1.29) | 0.367 |
| **Institution number** | 12 (4.4%) | 259 (95.6%) | 1 (1-2) | 1-9 | - | 1.20 (1.11-1.28) | **<0.001** | 256 | 1.18 (1.11-1.25) | **<0.0001** |
| **Institution type** | | | | | | | | | | |
| **Tertiary** | 19 (7%) | 252 (93%) | 144 (57.1%) | - | 12 (6-26) | *ref* | 0.586 | - | *ref* | 0.757 |
| **Hospital (unspecified)** | - | - | 12 (4.8%) | - | 8 (7-10) | 0.76 (0.38-1.49) | 0.420 | 247 | 0.85 (0.50-1.46) | 0.563 |
| **LTCF** | - | - | 2 (0.8%) | - | 64 (38-89) | 2.80 (0.57-13.72) | 0.202 | 247 | 1.36 (0.38-4.91) | 0.640 |
| **Network** | - | - | 88 (34.9%) | - | 9 (4-28) | 1.12 (0.76-1.65) | 0.563 | 247 | 1.18 (0.81-1.72) | 0.389 |
| **Secondary** | - | - | 6 (2.4%) | - | 11 (7-35) | 1.12 (0.44-2.87) | 0.809 | 247 | 0.77 (0.34-1.75) | 0.529 |
| **Institution capacity** | 174 (64.2%) | 97 (35.8%) | 1000 (700-1278) | 334-2420 | - | 1.00 (1.00-1.00) | 0.723 | 95 | 1.00 (1.00-1.00) | 0.748 |

| Variable / Category | Number (%) missing | Number (%) outbreaks with data available | Median (IQR) or Number (%) | Range | Median (IQR) number patients affected | Univariable Fold-change (95% CIs) | p-value† | Multi-variable N | Multivariable‡ Fold-change (95% CIs) | p-value† |
|---|---|---|---|---|---|---|---|---|---|---|
| **Ward number** | 112 (41.3%) | 159 (58.7%) | 2 (1-4) | 1-9 | - | 1.21 (1.13-1.29) | **<0.001** | 155 | 1.10 (1.05-1.18) | **<0.001** |
| **Ward types*** | | | | | | | | | | |
| **Medical ICU** | 103 (38%) | 168 (62%) | 96 (57.1%) | - | 12 (6-27) | 1.08 (0.76-1.53) | 0.677 | 163 | 1.06 (0.82-1.37) | 0.669 |
| **General surgical** | 103 (38%) | 168 (62%) | 39 (23.2%) | - | 18 (9-35) | 1.4 (0.94-2.07) | 0.095 | 163 | 1.25 (0.91-1.69) | 0.162 |
| **Neurology** | 103 (38%) | 168 (62%) | 32 (19%) | - | 11 (6-25) | 0.92 (0.59-1.41) | 0.691 | 163 | 1.02 (0.72-1.44) | 0.920 |
| **Neonatal ICU** | 103 (38%) | 168 (62%) | 27 (16.1%) | - | 19 (8-30) | 1.12 (0.71-1.77) | 0.627 | 163 | 1.11 (0.79-1.58) | 0.539 |
| **General medical** | 103 (38%) | 168 (62%) | 25 (14.9%) | - | 22 (14-44) | 1.56 (0.99-2.48) | 0.057 | 163 | 1.21 (0.84-1.74) | 0.300 |
| **Surgical ICU** | 103 (38%) | 168 (62%) | 21 (12.5%) | - | 13 (7-34) | 0.97 (0.59-1.61) | 0.915 | 163 | 0.90 (0.61-1.32) | 0.575 |
| **Respiratory** | 103 (38%) | 168 (62%) | 19 (11.3%) | - | 17 (8-54) | 1.64 (0.97-2.76) | 0.063 | 163 | 1.47 (0.96-2.25) | 0.079 |
| **Paediatric** | 103 (38%) | 168 (62%) | 17 (10.1%) | - | 20 (12-45) | 1.55 (0.89-2.68) | 0.119 | 163 | 1.61 (1.06-2.45) | **0.025** |
| **Cardiology** | 103 (38%) | 168 (62%) | 17 (10.1%) | - | 23 (8-66) | 1.76 (1.01-3.08) | **0.046** | 163 | 1.62 (1.07-2.45) | **0.023** |
| **Emergency department** | 103 (38%) | 168 (62%) | 16 (9.5%) | - | 32 (15-50) | 2.01 (1.16-3.48) | **0.013** | 163 | 1.95 (1.28-2.98) | **0.002** |
| **Infectious diseases department.** | 103 (38%) | 168 (62%) | 15 (8.9%) | - | 8 (5-21) | 0.8 (0.45-1.41) | 0.433 | 163 | 0.74 (0.47-1.15) | 0.175 |
| **Haematology** | 103 (38%) | 168 (62%) | 14 (8.3%) | - | 26 (7-43) | 1.38 (0.76-2.5) | 0.292 | 163 | 0.96 (0.58-1.59) | 0.881 |
| **Rehabilitation** | 103 (38%) | 168 (62%) | 12 (7.1%) | - | 6 (4-10) | 0.46 (0.23-0.91) | **0.027** | 163 | 0.62 (0.37-1.03) | 0.067 |
| **Gastroenterology / Hepatology** | 103 (38%) | 168 (62%) | 12 (7.1%) | - | 14 (9-21) | 1.07 (0.55-2.07) | 0.843 | 163 | 0.99 (0.60-1.62) | 0.960 |
| **Orthopaedics** | 103 (38%) | 168 (62%) | 11 (6.5%) | - | 14 (7-35) | 1.07 (0.54-2.12) | 0.834 | 163 | 0.92 (0.55-1.53) | 0.735 |
| **Urology** | 103 (38%) | 168 (62%) | 11 (6.5%) | - | 46 (32-56) | 2.78 (1.41-5.45) | **0.003** | 163 | 1.95 (1.13-3.37) | **0.016** |
| **Transplant** | 103 (38%) | 168 (62%) | 11 (6.5%) | - | 7 (6-14) | 0.64 (0.31-1.33) | 0.232 | 163 | 0.88 (0.52-1.46) | 0.611 |
| **Endemicity pre-outbreak*** | 75 (27.7%) | 196 (72.3%) | 102 (52%) | - | 10 (5-40) | 1.04 (0.73-1.49) | 0.814 | 193 | 1.24 (0.94-1.62) | 0.123 |
| **Background screening*** | 177 (65.3%) | 94 (34.7%) | 84 (89.4%) | - | 12 (6-27) | 0.6 (0.28-1.27) | 0.178 | 93 | 0.76 (0.44-1.31) | 0.312 |
| **Outbreak Detection method** | | | | | | | | | | |
| **clinical** | 118 (43.5%) | 153 (56.5%) | 74 (48.4%) | - | 10 (5-21) | *ref* | 0.017 | - | *ref* | 0.766 |
| **retrospective** | 118 (43.5%) | 153 (56.5%) | 25 (16.3%) | - | 6 (3-10) | 0.59 (0.31-1.13) | 0.110 | 150 | 1.10 (0.58-2.09) | 0.763 |
| **screening** | 118 (43.5%) | 153 (56.5%) | 54 (35.3%) | - | 14 (6-41) | 1.46 (0.98-2.17) | 0.061 | 150 | 1.13 (0.81-1.58) | 0.471 |

| Variable / Category | Number (%) missing | Number (%) outbreaks with data available | Median (IQR) or Number (%) | Range | Median (IQR) number patients affected | Univariable Fold-change (95% CIs) | p-value† | p-value (LRT)† | Multi-variable N | Multivariable‡ Fold-change (95% CIs) | p-value† |
|---|---|---|---|---|---|---|---|---|---|---|---|
| **Transmission mechanism*** | | | | | | | | | | | |
| **Clonal** | 0 (0%) | 271 (100%) | 253 (93.4%) | - | 10 (5-26) | 1.2 (0.69-2.08) | 0.518 | 0.517 | 256 | 1.82 (1.15-2.88) | **0.011** |
| **Plasmid** | 0 (0%) | 271 (100%) | 99 (36.5%) | - | 14 (7-41) | 1.78 (1.32-2.39) | **<0.001** | **<0.001** | 256 | 1.09 (0.80-1.47) | 0.588 |
| **Transposon / IS** | 0 (0%) | 271 (100%) | 19 (7%) | - | 27 (6-98) | 1.97 (1.16-3.33) | **0.012** | **0.011** | 256 | 1.12 (0.70-1.81) | 0.626 |
| **Outbreak definition*** | | | | | | | | | | | |
| **Epidemiological** | 114 (42.1%) | 157 (57.9%) | 41 (26.1%) | - | 9 (5-19) | 0.86 (0.58-1.27) | 0.440 | 0.433 | 153 | 0.94 (0.68-1.29) | 0.684 |
| **Clonal** | 114 (42.1%) | 157 (57.9%) | 104 (66.2%) | - | 9 (5-18) | 0.59 (0.42-0.84) | **0.004** | **0.003** | 153 | 0.86 (0.59-1.25) | 0.424 |
| **Plasmid** | 114 (42.1%) | 157 (57.9%) | 47 (29.9%) | - | 11 (6-25) | 1.29 (0.87-1.9) | 0.204 | 0.211 | 153 | 1.10 (0.79-1.53) | 0.576 |
| **Small MGE** | 114 (42.1%) | 157 (57.9%) | 3 (1.9%) | - | 20 (16-28) | 1.72 (0.4-7.4) | 0.454 | 0.439 | 153 | 0.57 (0.18-1.84) | 0.344 |
| **Gene-based** | 114 (42.1%) | 157 (57.9%) | 30 (19.1%) | - | 19 (8-40) | 2 (1.32-3.03) | **0.001** | **0.001** | 153 | 1.40 (0.95-2.04) | 0.085 |

* Compared to the absence of reporting of respective variables as the reference group.

†p-values are for a Wald test. p-values appearing in the reference ('ref') category row represent overall Wald-test p-values for that category, obtained using Satterthwaite's method for a type III ANOVA for mixed effects models. p-values <0.05 are shown in **bold**.

‡ Multivariable associations are adjusted for all 6 variables in the final core model: number of outbreaks reported in study, publication year, number of institutions, start year, number of STs involved, clonal transmission involved.

Abbreviations; CIs – confidence intervals; IS – insertion sequence; IQR- interquartile range.

**Table S5: Genomic and microbiological risk/protective factors for outbreak size with descriptive summary, univariable and multivariable associations with outbreak size (total number of patients infected/colonised per outbreak).** Descriptive summaries are provided considering the dataset of 271 outbreaks with complete outcome data (out of 272 included in this review). Univariable associations are calculated from a dataset of outbreaks with complete data for each variable (shown in the 3rd column from the left). Multivariable associations are calculated from a dataset of outbreaks with complete data on that variable as well as all of the variables included in the final model. Estimates and 95% confidence intervals (CIs) are reported on the original (non-$log_{10}$) scale, interpreted as relative changes in outbreak size (fold-change in size).

| **Variable** / **Category** | **Number (%) missing** | **Number (%) outbreaks with data available** | **Median (IQR) or number (%)** | **Range** | **Median (IQR) number patients affected** | **Univariable Fold-change (95% CIs)** | **p-value†** | **Multi-variable N** | **Multivariable‡ Fold-change (95% CIs)** | **p-value†** |
|---|---|---|---|---|---|---|---|---|---|---|
| **Number of Enterobacterales spp.** | 0 (0%) | 271 (100%) | 1 (1-2) | 1-5 | - | 1.33 (1.21-1.47) | **<0.001** | 256 | 1.02 (0.86-1.2) | 0.812 |
| **Main Enterobacterales spp.** | | | | | | | | | | |
| ***K. pneumoniae* complex** | 0 (0%) | 271 (100%) | 171 (63.1%) | - | 10 (5-28) | *ref* | 0.903 | 256 | *ref* | 0.390 |
| ***Citrobacter* spp.** | - | - | 13 (4.8%) | - | 8 (6-12) | 0.73 (0.39-1.37) | 0.324 | 256 | 0.68 (0.41-1.15) | 0.151 |
| ***E. cloacae* complex** | - | - | 48 (17.7%) | - | 11 (6-28) | 1.00 (0.67-1.48) | 0.985 | 256 | 0.82 (0.59-1.15) | 0.246 |
| ***E. coli*** | - | - | 22 (8.1%) | - | 6 (3-28) | 0.94 (0.58-1.55) | 0.821 | 256 | 0.84 (0.56-1.28) | 0.416 |
| **Other** | - | - | 17 (6.3%) | - | 14 (7-19) | 0.92 (0.53-1.61) | 0.778 | 256 | 0.73 (0.45-1.16) | 0.183 |
| **Number of Enterobacterales STs** | 0 (0%) | 271 (100%) | 1 (1-3) | 1-12 | - | 1.16 (1.11-1.20) | **<0.001** | 256 | 1.14 (1.10-1.18) | **<0.001** |
| **Main Enterobacterales STs*** | | | | | | | | | | |
| ***Other K. pneumoniae STs*** | 8 (3%) | 263 (97%) | 75 (28.5%) | - | 9 (4-19) | *ref* | 0.667 | 249 | *ref* | 0.700 |
| ***Citrobacter freundii STs*** | - | - | 9 (3.4%) | - | 8 (7-12) | 1.02 (0.47-2.20) | 0.964 | 249 | 0.92 (0.49-1.75) | 0.807 |
| ***E. cloacae* complex STs** | - | - | 48 (18.3%) | - | 12 (6-33) | 1.21 (0.78-1.89) | 0.396 | 249 | 0.97 (0.67-1.41) | 0.868 |
| ***E. coli STs*** | - | - | 18 (6.8%) | - | 7 (3-33) | 1.18 (0.66-2.09) | 0.574 | 249 | 1.13 (0.70-1.83) | 0.607 |
| **ST11 *K. pneumoniae*** | - | - | 34 (12.9%) | - | 9 (5-42) | 1.18 (0.75-1.86) | 0.481 | 249 | 1.33 (0.90-1.97) | 0.152 |
| **ST14 *K. pneumoniae*** | - | - | 17 (6.5%) | - | 9 (5-48) | 1.60 (0.89-2.88) | 0.119 | 249 | 1.32 (0.80-2.18) | 0.269 |
| **ST15 *K. pneumoniae*** | - | - | 16 (6.1%) | - | 10 (5-49) | 1.24 (0.68-2.26) | 0.480 | 249 | 1.36 (0.79-2.35) | 0.264 |
| **ST258 *K. pneumoniae*** | - | - | 18 (6.8%) | - | 12 (6-20) | 1.19 (0.65-2.20) | 0.569 | 249 | 1.20 (0.73-1.96) | 0.468 |
| **ST307 *K. pneumoniae*** | - | - | 10 (3.8%) | - | 36 (12-58) | 2.17 (1.01-4.67) | 0.048 | 249 | 1.32 (0.71-2.47) | 0.382 |
| **Other STs** | - | - | 18 (6.8%) | - | 14 (6-19) | 0.91 (0.51-1.62) | 0.749 | 249 | 0.83 (0.50-1.38) | 0.481 |
| **Non-Enterobacterales spp. present vs absent**** | 0 (0%) | 271 (100%) | 4 (1.5%) | - | 8 (6-19) | 0.80 (0.26-2.47) | 0.697 | 256 | 0.46 (0.16-1.29) | 0.139 |
| **Main Carbapenemase allele** | | | | | | | | | | |
| **NDM-1** | 0 (0%) | 271 (100%) | 56 (20.7%) | - | 9 (5-25) | *ref* | 0.332 | 256 | *ref* | 0.134 |
| **Other IMP** | - | - | 5 (1.8%) | - | 11 (3-18) | 0.70 (0.26-1.86) | 0.468 | 256 | 0.67 (0.30-1.52) | 0.340 |
| **IMP-4** | - | - | 10 (3.7%) | - | 20 (6-84) | 1.13 (0.52-2.47) | 0.760 | 256 | 0.84 (0.42-1.68) | 0.622 |

| Variable / Category | Number (%) missing | Number (%) outbreaks with data available | Median (IQR) or number (%) | Range | Median (IQR) number patients affected | Univariable Fold-change (95% CIs) | p-value† | Multi-variable N | Multivariable‡ Fold-change (95% CIs) | p-value† |
|---|---|---|---|---|---|---|---|---|---|---|
| **KPC-2** | - | - | 53 (19.6%) | - | 11 (6-30) | 1.06 (0.68-1.65) | 0.793 | 256 | 1.12 (0.78-1.60) | 0.538 |
| **KPC-3** | - | - | 21 (7.7%) | - | 7 (4-20) | 0.68 (0.38-1.22) | 0.195 | 256 | 0.93 (0.58-1.51) | 0.780 |
| **Mixed** | - | - | 22 (8.1%) | - | 22 (6-42) | 1.31 (0.76-2.24) | 0.330 | 256 | 1.28 (0.81-2.01) | 0.285 |
| **Other NDM** | - | - | 20 (7.4%) | - | 8 (3-16) | 0.72 (0.41-1.26) | 0.250 | 256 | 0.75 (0.46-1.21) | 0.232 |
| **OXA-48** | - | - | 33 (12.2%) | - | 10 (5-41) | 0.99 (0.60-1.61) | 0.953 | 256 | 1.33 (0.88-2.02) | 0.179 |
| **Other OXA-48-like** | - | - | 18 (6.6%) | - | 16 (4-54) | 1.54 (0.85-2.8) | 0.154 | 256 | 1.63 (0.99-2.68) | 0.054 |
| **Other VIM** | - | - | 8 (3%) | - | 8 (7-12) | 0.61 (0.25-1.49) | 0.277 | 256 | 0.63 (0.30-1.33) | 0.226 |
| **VIM-1** | - | - | 16 (5.9%) | - | 11 (6-22) | 0.75 (0.39-1.45) | 0.391 | 256 | 0.76 (0.45-1.30) | 0.316 |
| **Main Carbapenemase gene family** | | | | | | | | | | |
| **KPC** | 0 (0%) | 271 (100%) | 83 (30.6%) | - | 10 (5-22) | *ref* | 0.374 | 256 | *ref* | 0.064 |
| **IMP** | - | - | 16 (5.9%) | - | 14 (6-55) | 1.18 (0.64-2.17) | 0.594 | 256 | 0.77 (0.45-1.31) | 0.335 |
| **NDM** | - | - | 78 (28.8%) | - | 9 (4-19) | 1.02 (0.71-1.47) | 0.917 | 256 | 0.89 (0.66-1.21) | 0.456 |
| **OXA-48** | - | - | 33 (12.2%) | - | 10 (5-41) | 1.11 (0.70-1.77) | 0.645 | 256 | 1.28 (0.87-1.90) | 0.209 |
| **OXA-48-like** | - | - | 18 (6.6%) | - | 16 (4-54) | 1.77 (0.99-3.15) | 0.054 | 256 | 1.57 (0.97-2.56) | 0.068 |
| **VIM** | - | - | 24 (8.9%) | - | 10 (6-16) | 0.80 (0.44-1.45) | 0.469 | 256 | 0.69 (0.42-1.13) | 0.139 |
| **Mixed** | - | - | 19 (7%) | - | 23 (6-42) | 1.40 (0.80-2.45) | 0.237 | 256 | 1.20 (0.75-1.90) | 0.443 |
| **Number of carbapenemase alleles** | 0 (0%) | 271 (100%) | 1 (1-1) | 1-2 | - | 1.66 (1.17-2.36) | **0.005** | 256 | 1.24 (0.91-1.70) | 0.170 |
| **Number of carbapenemase-assoc. plasmids** | 46 (17%) | 225 (83%) | 1 (1-2) | 1-4 | - | 1.40 (1.16-1.68) | **<0.001** | 219 | 1.21 (1.02-1.43) | **0.025** |
| **Number of replicon types per isolate** | 113 (41.7%) | 158 (58.3%) | 3 (2-5) | 1-10 | - | 1.12 (1.03-1.22) | **0.009** | 153 | 1.01 (0.93-1.09) | 0.844 |
| **Number other MGEs per isolate** | 144 (53.1%) | 127 (46.9%) | 1 (1-2) | 0-6 | - | 1.18 (1.03-1.35) | **0.018** | 123 | 1.12 (0.99-1.27) | 0.074 |
| **Small MGE types involved** | | | | | | | | | | |
| **Any Integron** | 148 (54.6%) | 123 (45.4%) | 28 (22.8%) | - | 18 (8-41) | 1.29 (0.75-2.21) | 0.351 | 119 | 1.11 (0.72-1.72) | 0.619 |
| **Any Transposon** | - | - | 71 (57.7%) | - | 20 (8-44) | 1.62 (1.04-2.52) | **0.034** | 119 | 1.21 (0.83-1.77) | 0.324 |
| **Any Insertion Sequence** | - | - | 31 (25.2%) | - | 12 (6-40) | 0.80 (0.50-1.27) | 0.345 | 119 | 0.78 (0.53-1.15) | 0.209 |
| **Specific small MGEs involved** | | | | | | | | | | |
| **Tn4401** | - | - | 30 (24.4%) | - | 18 (5-36) | 0.93 (0.57-1.52) | 0.765 | 119 | 0.80 (0.51-1.25) | 0.322 |
| **Tn125-like** | - | - | 22 (17.9%) | - | 19 (5-42) | 1.09 (0.61-1.94) | 0.763 | 119 | 1.10 (0.70-1.72) | 0.675 |
| **Tn3-like** | - | - | 11 (8.9%) | - | 24 (10-46) | 1.21 (0.60-2.41) | 0.591 | 119 | 0.91 (0.51-1.63) | 0.755 |
| **Other Transposon** | - | - | 17 (13.8%) | - | 19 (10-57) | 1.60 (0.90-2.83) | 0.108 | 119 | 1.50 (0.94-2.40) | 0.092 |

| Variable Category | Number (%) missing | Number (%) outbreaks with data available | Median (IQR) or number (%) | Range | Median (IQR) number patients affected | Univariable Fold-change (95% CIs) | p-value† | Multi-variable N | Multivariable‡ Fold-change (95% CIs) | p-value† |
|---|---|---|---|---|---|---|---|---|---|---|
| **Other IS** | - | - | 26 (21.1%) | - | 10 (6-30) | 0.65 (0.40-1.07) | 0.090 | 119 | 0.75 (0.50-1.13) | 0.167 |
| **Number of phenotypic resistances** | 204 (75.3%) | 67 (24.7%) | 11 (7-14) | 4-19 | - | 1.00 (0.94-1.06) | 0.974 | 66 | 0.99 (0.95-1.04) | 0.738 |
| **Number of ARGs per isolate** | 105 (38.7%) | 166 (61.3%) | 14 (9-18) | 3-25 | - | 1.03 (1.01-1.06) | **0.016** | 161 | 1.02 (0.99-1.04) | 0.136 |
| **Number of VF genes per isolate** | 183 (67.5%) | 88 (32.5%) | 3 (1-6) | 0-15 | - | 1.02 (0.97-1.08) | 0.442 | 85 | 1.03 (0.99-1.08) | 0.153 |
| **Main plasmid identity** | | | | | | | | | | |
| **pOXA-48**** | 63 (23.2%) | 208 (76.8%) | 11 (5.3%) | - | 19 (9-42) | 1.68 (0.88-3.21) | 0.118 | 203 | 1.51 (0.86-2.66) | 0.149 |
| **Main plasmid Inc type** | | | | | | | | | | |
| **IncFII** | 87 (32.1%) | 184 (67.9%) | 22 (12%) | - | 12 (6-50) | *ref* | 0.241 | 179 | ref | 0.422 |
| **IncFIB** | 87 (32.1%) | 184 (67.9%) | 12 (6.5%) | - | 7 (5-10) | 0.57 (0.25-1.28) | 0.172 | 179 | 0.85 (0.45-1.62) | 0.617 |
| **IncL** | 87 (32.1%) | 184 (67.9%) | 10 (5.4%) | - | 14 (9-30) | 0.98 (0.41-2.34) | 0.968 | 179 | 1.06 (0.53-2.13) | 0.870 |
| **IncL/M** | 87 (32.1%) | 184 (67.9%) | 14 (7.6%) | - | 36 (12-44) | 2.02 (0.91-4.5) | 0.085 | 179 | 1.67 (0.90-3.10) | 0.101 |
| **IncN** | 87 (32.1%) | 184 (67.9%) | 21 (11.4%) | - | 7 (6-26) | 0.76 (0.38-1.51) | 0.428 | 179 | 0.80 (0.46-1.39) | 0.425 |
| **IncX** | 87 (32.1%) | 184 (67.9%) | 22 (12%) | - | 11 (6-24) | 0.96 (0.49-1.90) | 0.912 | 179 | 1.00 (0.59-1.69) | 0.987 |
| **Multireplicon** | 87 (32.1%) | 184 (67.9%) | 36 (19.6%) | - | 12 (7-31) | 1.09 (0.61-1.97) | 0.766 | 179 | 0.89 (0.55-1.43) | 0.623 |
| **Other single replicon** | 87 (32.1%) | 184 (67.9%) | 47 (25.5%) | - | 14 (6-38) | 1.05 (0.58-1.89) | 0.869 | 179 | 0.90 (0.57-1.41) | 0.638 |
| **Main plasmid size (kbp)** | 120 (44.3%) | 151 (55.7%) | 0.68 (0.53-1.34) | 0.36-2.78 | - | 1.00 (0.77-1.30) | 0.985 | 149 | 0.94 (0.76-1.18) | 0.614 |
| **Number of ARGs on main plasmid** | 168 (62%) | 103 (38%) | 5 (2-9) | 1-13 | - | 0.98 (0.93-1.03) | 0.411 | 100 | **0.95 (0.91-0.99)** | **0.013** |
| **Number of small MGEs on main plasmid** | 199 (73.4%) | 72 (26.6%) | 1 (1-2) | 1-10 | - | 0.93 (0.82-1.06) | 0.301 | 71 | 0.93 (0.83-1.03) | 0.151 |
| **Plasmid conjugation ability**** | | | | | | | | | | |
| **non-conjugative vs conj.** | 212 (78.2%) | 59 (21.8%) | 13 (22%) | - | 12 (5-26) | 0.70 (0.33-1.5) | 0.351 | 59 | 0.86 (0.45-1.65) | 0.640 |

*The ‘Other *K. pneumoniae* STs’ category was the reference category for the Main ST variable, and comprised eight outbreaks with *K. pneumoniae* ST437, sever ST101, six ST51, five ST16, three each of ST17, ST405, ST78 and unspecified STs, two each of ST20 and ST54, and one each of ST1026, ST104, ST196, ST219, ST2253, ST23, ST231, ST2407, ST2497, ST25, ST26, ST283, ST2084, ST34, ST340, ST348, ST37, ST383, ST39, ST392, ST395, ST392, ST395, ST476, ST496, ST502, ST526, ST5551, ST571, ST661, ST716, ST76, ST859, ST873, ST968. The ‘*Escherichia coli* STs’ category comprised five outbreaks with ST410, three ST313, 2 ST167, and one each of ST216, ST378, ST38, ST399, ST405, ST6388, and ST648, which were grouped together for statistical power. The ‘Other STs’ included three outbreaks with *Serratia marcescens* as the main Enterobacterales, two each of *Citrobacter farmerii* and *Klebsiella oxytoca* ST14, and one each of *Leclercia adenocarboxylata, Morganella morganii, Mixta calida, Proteus mirabilis, Salmonella* Goldcoast ST358, *Klebsiella variicola* ST15, *Klebsiella variicola* ST3456, *Klebsiella oxytoca* ST220, *Klebsiella oxytoca* ST29, and *Klebsiella aerogenes* ST233. The ‘*Enterobacter cloacae* complex STs’ comprised six outbreaks ST89, four unspecified STs, three each of ST11, ST66, ST78, ST90, two of ST121, ST171, ST177, ST182, ST93, and one each of ST1015, ST1283, ST1303, ST131, ST134, ST14, ST190, ST198, ST269, ST310, ST428, ST595, ST69, ST873, ST94, and ST97. The ‘*Citrobacter freundii* STs’ comprised three outbreaks ST18, two each of ST22 and ST65, and one each of ST523, ST98, and one unspecified ST.

** Compared to reference group of ‘absent’ other Enterobacterales spp., or non-pOXA-48 plasmids for Main plasmid identity; associations are compared to the absence of reporting of respective variables as the reference group.

†p-values are for a Wald test. p-values appearing in the reference (‘ref’) category row represent overall Wald-test p-values for that category, obtained using Satterthwaite's method for a type III ANOVA for mixed effects models. p-values <0.05 are shown in **bold**.

‡ Multivariable associations are adjusted for all 6 variables in the final core model: number of outbreaks reported in study, publication year, number of institutions, start year, number of STs involved, clonal transmission involved. Abbreviations: ARG – antimicrobial resistance genes; CIs – confidence intervals; conj. – conjugative; IQR – interquartile range; IS – insertion sequence; ; kbp – kilobase-pair; MGE – mobile genetic element; ST – multi-locus sequence type; VF – virulence factor.

**Table S6: Infection control measures reported with descriptive summary, univariate and multivariate associations with outbreak size (total number of patients infected/colonised per outbreak).** Descriptive summaries are provided considering the dataset of 271 outbreaks with complete outcome data (out of 272 included in this review). Univariable associations are calculated from a dataset of outbreaks with complete data for each variable (shown in the 3rd column from the left). Multivariable associations are calculated from a dataset of outbreaks with complete data on that variable as well as all of the variables included in the final model. Estimates and 95% confidence intervals (CIs) are reported on the original (non-$\log_{10}$) scale, interpreted as relative changes in outbreak size (fold-change in size).

| Variable | Number (%) missing | Number (%) of outbreaks with data available | Median (IQR) or Number %) | Range | Median (IQR) or number of patients affected | Univariate Fold-change (95% CIs) | p-value† | Multi-variate N | Multivariate‡ Fold-change (95% CIs) | p-value† |
|---|---|---|---|---|---|---|---|---|---|---|
| **Any IPC measure reported** | 0 (0%) | 271 (100%) | 112 (41.3%) | - | 12 (6-29) | 1.16 (0.85-1.58) | 0.343 | 256 | 0.86 (0.66-1.13) | 0.282 |
| **Any Patient IPC measure** | 159 (58.7%) | 112 (41.3%) | 97 (86.6%) | - | 12 (5-29) | 0.92 (0.48-1.78) | 0.808 | 109 | 0.94 (0.56-1.60) | 0.821 |
| **Enhanced screening** | - | - | 31 (27.7%) | - | 23 (10-56) | 2.13 (1.36-3.34) | **0.001** | 109 | 1.83 (1.28-2.61) | **0.001** |
| **Patient cohorting** | - | - | 26 (23.2%) | - | 22 (9-48) | 1.75 (1.07-2.87) | **0.026** | 109 | 1.46 (1.00-2.14) | 0.052 |
| **Contact precautions** | - | - | 61 (54.5%) | - | 12 (5-40) | 1.00 (0.64-1.55) | 0.998 | 109 | 1.00 (0.71-1.40) | 0.982 |
| **Screening during hospitalisation** | - | - | 48 (42.9%) | - | 18 (7-38) | 1.23 (0.79-1.91) | 0.357 | 109 | 1.03 (0.73-1.46) | 0.848 |
| **Contact tracing** | - | - | 39 (34.8%) | - | 7 (4-28) | 0.68 (0.43-1.08) | 0.105 | 109 | 0.84 (0.59-1.20) | 0.337 |
| **–Other patient IPC intervention** | - | - | 30 (26.8%) | - | 20 (8-41) | 1.60 (1.00-2.56) | 0.052 | 109 | 1.37 (0.95-1.97) | 0.093 |
| **Any HCW IPC measure** | 159 (58.7%) | 112 (41.3%) | 77 (68.8%) | - | 15 (7-36) | 1.34 (0.82-2.18) | 0.241 | 109 | 1.23 (0.84-1.81) | 0.283 |
| **HCW Screening** | - | - | 15 (13.4%) | - | 26 (8-56) | 1.89 (1.02-3.50) | **0.042** | 109 | 1.73 (1.08-2.76) | **0.022** |
| **HCW Education** | - | - | 29 (25.9%) | - | 20 (10-47) | 1.89 (1.18-3.02) | **0.008** | 109 | 1.32 (0.90-1.94) | 0.153 |
| **HCW Enhanced hand hygiene** | - | - | 30 (26.8%) | - | 14 (6-24) | 1.01 (0.62-1.65) | 0.96 | 109 | 1.17 (0.81-1.70) | 0.403 |
| **HCW cohorting** | - | - | 25 (22.3%) | - | 9 (5-41) | 1.14 (0.66-1.96) | 0.631 | 109 | 1.08 (0.73-1.58) | 0.709 |
| **Outbreak response team** | - | - | 16 (14.3%) | - | 12 (8-24) | 1.12 (0.60-2.09) | 0.727 | 109 | 0.98 (0.62-1.57) | 0.948 |
| **Other HCW IPC intervention** | - | - | 45 (40.2%) | - | 18 (7-40) | 1.34 (0.87-2.08) | 0.182 | 109 | 1.18 (0.82-1.70) | 0.373 |
| **Any Environmental IPC measure** | 159 (58.7%) | 112 (41.3%) | 85 (75.9%) | - | 14 (7-40) | 1.52 (0.90-2.55) | 0.113 | 109 | 1.38 (0.92-2.06) | 0.120 |
| **Environmental Screening** | - | - | 62 (55.4%) | - | 14 (7-37) | 1.14 (0.74-1.76) | 0.545 | 109 | 1.09 (0.78-1.53) | 0.598 |
| **Environmental Cleaning** | - | - | 59 (52.7%) | - | 12 (7-40) | 1.21 (0.78-1.88) | 0.384 | 109 | 1.20 (0.84-1.70) | 0.313 |
| **Equipment Replacement** | - | - | 24 (21.4%) | - | 16 (7-42) | 1.13 (0.67-1.93) | 0.641 | 109 | 1.22 (0.79-1.88) | 0.358 |
| **Ward closure** | - | - | 13 (11.6%) | - | 20 (7-66) | 1.50 (0.78-2.87) | 0.217 | 109 | 1.36 (0.81-2.27) | 0.242 |
| **Other environmental intervention** | - | - | 11 (9.8%) | - | 40 (8-54) | 2.01 (0.94-4.31) | 0.073 | 109 | 1.86 (1.07-3.21) | **0.028** |

* Compared to not reporting that specific control measure, among outbreaks reporting any control measures.

†p-values are for a Wald test. p-values appearing in the reference ('ref') category row represent overall Wald-test p-values for that category, obtained using Satterthwaite's method for a type III ANOVA for mixed effects models. p-values <0.05 are shown in **bold**.

‡ Multivariate associations are adjusted for all 6 variables in the final core model: no outbreaks reported in study, publication year, number of institutions, start year, number of STs involved, clonal transmission involved. Abbreviations: IQR- interquartile range; CIs – confidence intervals; HCW- healthcare worker; IPC – infection prevention and control.

**Table S7: Descriptive summary, univariate and multivariate associations between variables included in the final backwards-selected multivariate mixed effect linear regression model for $log_{10}$(number of patients affected per outbreak), for the subset of outbreaks affecting ≥10 or more patients ('10-or-more-patients' model).** Descriptive summaries are provided considering the dataset of 140 outbreaks (out of 272 included in this review) reported to affect ≥10 patients. Univariate associations are calculated using the outbreaks with complete data for each variable (shown in the 3rd column from the left). Multivariate associations are calculated from a dataset of 133 outbreaks with complete data on the all of the variables included in the final model. Estimates and 95% confidence intervals (CIs) are reported on the original (non-$log_{10}$) scale, interpreted as relative changes in outbreak size (fold-change in size). This model had a marginal $R^2$ of 0.334 (conditional $R^2$ could not be evaluated due to data sparsity), AIC 65.2, and BIC of 114.3.

| Variable | Category | Number (%) missing | Number (%) outbreaks with data available | Median (IQR) or Number (%) | Range | Median (IQR) or number patients affected | Univariable Fold-change (95% CIs) | p-value† | Multivariable Fold-change (95% CIs) | p-value† |
|---|---|---|---|---|---|---|---|---|---|---|
| **Publication type** | **Journal article** | 0 (0%) | 140 (100%) | 124 (88.6%) | - | 26 (14-47) | *ref* | 0.714 | - | **0.012** |
| | **Abstract** | - | - | 13 (9.3%) | - | 43 (16-72) | 1.17 (0.71-1.94) | 0.521 | 1.97 (1.22-3.17) | **0.006** |
| | **Preprint** | - | - | 3 (2.1%) | - | 15 (13-46) | 0.81 (0.33-1.98) | 0.634 | 0.66 (0.31-1.39) | 0.268 |
| **Number of outbreaks** | | 0 (0%) | 140 (100%) | 1 (1-2) | 1-9 | - | 0.98 (0.91-1.05) | 0.538 | 0.93 (0.87-0.99) | **0.031** |
| **Transposon/IS** | **Not involved** | 0 (0%) | 140 (100%) | 127 (90.7%) | - | 25 (14-47) | *ref* | ref | - | **-** |
| | **Involved in transmission** | - | - | 13 (9.3%) | - | 65 (27-100) | 1.94 (1.26-2.98) | 0.003 | 1.53 (1.03-2.26) | **0.035** |
| **Institution number** | | 3 (2.1%) | 137 (97.9%) | 1 (1-3) | 1-9 | - | 1.08 (1.02-1.15) | 0.005 | 1.10 (1.04-1.16) | **0.001** |
| **Number Enterobacterales STs** | | 0 (0%) | 140 (100%) | 1 (1-5.25) | 1-10 | - | 1.05 (1.02-1.09) | 0.002 | 1.04 (1.01-1.08) | **0.011** |
| **Number. Carbapenemase alleles involved** | | 0 (0%) | 140 (100%) | 1 (1-1) | 1-2 | - | 1.73 (1.27-2.36) | 0.001 | 1.30 (0.96-1.77) | 0.094 |
| **Main Enterobacterales STs*** | | | | | | | | | | |
| | ***Other K. pneumoniae STs*** | 4 (2.9%) | 136 (97.1%) | 36 (26.5%) | - | 20 (14-31) | *ref* | 0.007 | - | **0.004** |
| | ***Citrobacter freundii STs*** | - | - | 3 (2.2%) | - | 65 (38-69) | 1.66 (0.69-3.97) | 0.251 | 1.25 (0.56-2.77) | 0.585 |
| | ***E. cloacae* complex STs** | - | - | 29 (21.3%) | - | 27 (12-48) | 1.28 (0.88-1.84) | 0.190 | 1.07 (0.76-1.50) | 0.698 |
| | ***E. coli STs*** | - | - | 7 (5.1%) | - | 66 (32-75) | 2.32 (1.28-4.18) | 0.006 | 2.13 (1.28-3.56) | **0.004** |
| | **ST11 *K. pneumoniae*** | - | - | 17 (12.5%) | - | 42 (21-49) | 1.46 (0.95-2.22) | 0.081 | 1.66 (1.13-2.42) | **0.010** |
| | **ST14 *K. pneumoniae*** | - | - | 8 (5.9%) | - | 62 (36-116) | 2.37 (1.35-4.15) | 0.003 | 1.82 (1.08-3.07) | **0.024** |
| | **ST15 *K. pneumoniae*** | - | - | 8 (5.9%) | - | 53 (23-76) | 1.73 (0.99-3.03) | 0.056 | 1.31 (0.81-2.13) | 0.273 |
| | **ST258 *K. pneumoniae*** | - | - | 10 (7.4%) | - | 20 (18-40) | 1.07 (0.64-1.79) | 0.803 | 1.01 (0.65-1.56) | 0.980 |
| | **ST307 *K. pneumoniae*** | - | - | 7 (5.1%) | - | 47 (36-61) | 2.11 (1.17-3.83) | 0.014 | 1.22 (0.72-2.08) | 0.458 |
| | **Other STs** | - | - | 11 (8.1%) | - | 19 (14-23) | 0.85 (0.51-1.39) | 0.509 | 0.72 (0.47-1.11) | 0.134 |

*The 'Other *K. pneumoniae* STs' category was the reference category for the Main ST variable, and comprised eight outbreaks with *K. pneumoniae* ST437, sever ST101, six ST51, five ST16, three each of ST17, ST405, ST78 and unspecified STs, two each of ST20 and ST54, and one each of ST1026, ST104, ST196, ST219, ST2253, ST23, ST231, ST2407, ST2497, ST25, ST26, ST283, ST2084, ST34, ST340, ST348, ST37, ST383, ST39, ST392, ST395, ST392, ST395, ST476, ST496, ST502, ST526, ST5551, ST571, ST661, ST716, ST76, ST859, ST873, ST968. The '*Escherichia coli* STs' category comprised five outbreaks with ST410, three ST313, 2 ST167, and one each of ST216, ST378, ST38, ST399, ST405, ST6388, and ST648, which were grouped together for statistical power. The 'Other STs' included three outbreaks with *Serratia marcescens* as the main Enterobacterales, two each of *Citrobacter farmerii* and *Klebsiella oxytoca* ST14, and one each of *Leclercia adenocarboxylata, Morganella morganii, Mixta calida, Proteus mirabilis, Salmonella* Goldcoast ST358, *Klebsiella variicola* ST15, *Klebsiella variicola* ST3456, *Klebsiella oxytoca* ST220, *Klebsiella oxytoca* ST29, and *Klebsiella aerogenes* ST233. The '*Enterobacter cloacae* complex STs' comprised six outbreaks ST89, four unspecified STs, three each of ST11, ST66, ST78, ST90, two of ST121, ST171, ST177, ST182, ST93, and one each of ST1015, ST1283, ST1303, ST131, ST134, ST14, ST190, ST198, ST269, ST310, ST428, ST595, ST69, ST873, ST94, and ST97. The '*Citrobacter freundii* STs' comprised three outbreaks ST18, two each of ST22 and ST65, and one each of ST523, ST98, and one unspecified ST.

[†]p-values are for a Wald test. p-values appearing in the reference (‘ref’) category row represent overall Wald-test p-values for that category, obtained using Satterthwaite's method for a type III ANOVA for mixed effects models. p-values <0.05 are shown in **bold**.
Abbreviations: IQR- interquartile range; CIs – confidence intervals; ST- Multi-locus sequence type.

**Table S8: Study-level risk/protective factors for outbreak size for the subset of outbreak affecting at least 10 patients, with descriptive summaries, univariable and multivariable associations with outbreak size (total number of patients infected/colonised per outbreak) ('10-or-more-patient' outbreaks model).** Descriptive summaries are provided considering the dataset of 140 outbreaks (out of 272 included in this review). Univariable associations are calculated from a dataset of outbreaks with complete data for each variable (shown in the 3rd column from the left). Multivariable associations are calculated from a dataset of outbreaks with complete data on that variable as well as all of the variables included in the final '10-or-more-patient' outbreaks model (publication type, number of outbreaks reported in study, transposon/IS involved in transmission, number of institutions, number of Enterobacterales STs involved, number of carbapenemase alleles involved, main Enterobacterales ST involved). Estimates and 95% confidence intervals (CIs) are reported on the original (non-$log_{10}$) scale, interpreted as relative changes in outbreak size (fold-change in size).

| Variable / Category | Number (%) missing | Number (%) of outbreaks with data available | Median (IQR) or number (%) | Range | Median (IQR) of number patients affected | Univariable Fold-change (95% CIs) | p-value† | Multi-variate N | Multivariable‡ Fold-change (95% CIs) | p-value† |
|---|---|---|---|---|---|---|---|---|---|---|
| **Publication year** | 0 (0%) | 140 (100%) | 2021 (2019-2022.25) | 2016-2023 | - | 1.01 (0.95-1.07) | 0.722 | 133 | 1.01 (0.96-1.06) | 0.613 |
| **Publication type** | | | | | | | | | | |
| **Journal article** | 0 (0%) | 140 (100%) | 124 (88.6%) | - | 26 (14-47) | *ref* | 0.714 | 133 | *ref* | **0.012** |
| **Abstract** | - | - | 13 (9.3%) | - | 43 (16-72) | 1.17 (0.71-1.94) | 0.521 | 133 | 1.97 (1.22-3.17) | **0.006** |
| **Preprint** | - | - | 3 (2.1%) | - | 15 (13-46) | 0.81 (0.33-1.98) | 0.634 | 133 | 0.66 (0.31-1.39) | 0.268 |
| **Study design** | | | | | | | | | | |
| **Outbreak report** | 0 (0%) | 140 (100%) | 71 (50.7%) | - | 25 (17-47) | *ref* | 0.477 | 133 | *ref* | 0.758 |
| **Case-control** | - | - | 2 (1.4%) | - | 27 (19-35) | 0.76 (0.22-2.56) | 0.645 | 133 | 0.60 (0.15-2.30) | 0.451 |
| **Cross-sectional** | - | - | 1 (0.7%) | - | 115 (115-115) | 4.00 (0.86-18.66) | 0.078 | 133 | 1.62 (0.44-5.98) | 0.463 |
| **Prospective descriptive** | - | - | 3 (2.1%) | - | 34 (28-46) | 1.20 (0.48-2.95) | 0.696 | 133 | 1.30 (0.62-2.71) | 0.479 |
| **Retrospective descriptive** | - | - | 63 (45%) | - | 27 (13-55) | 1.02 (0.77-1.35) | 0.884 | 133 | 0.95 (0.73-1.23) | 0.686 |
| **Funding** | | | | | | | | | | |
| **Government funding*** | 21 (15%) | 119 (85%) | 103 (86.6%) | - | 26 (14-48) | 0.87 (0.58-1.32) | 0.511 | 116 | 0.68 (0.47-0.98) | **0.041** |
| **Academic funding*** | - | - | 25 (21%) | - | 34 (20-44) | 1.16 (0.82-1.64) | 0.404 | 116 | 0.97 (0.71-1.32) | 0.833 |
| **Other funding*** | - | - | 19 (16%) | - | 33 (20-66) | 1.33 (0.90-1.95) | 0.147 | 116 | 1.33 (0.95-1.86) | 0.095 |
| **Conflict of interest** | | | | | | | | | | |
| **Declared present**** | 28 (20%) | 112 (80%) | 16 (14.3%) | - | 32 (17-56) | 1.2 (0.78-1.87) | 0.402 | 107 | 1.12 (0.79-1.60) | 0.516 |
| **Number of outbreaks** | 0 (0%) | 140 (100%) | 1 (1-2) | 1-9 | - | 0.98 (0.91-1.05) | 0.538 | 133 | 0.93 (0.87-0.99) | **0.031** |

* Compared to the absence of reporting that type of funding or conflict of interest, for those outbreaks where funding and conflict of interest was reported, respectively.

†p-values are for a Wald test. p-values appearing in the reference ('ref') category row represent overall Wald-test p-values for that category, obtained using Satterthwaite's method for a type III ANOVA for mixed effects models. p-values <0.05 are shown in **bold**.

‡ Multivariable associations are adjusted for all 6 variables in the final core model: number outbreaks reported in study, publication year, number of institutions, start year, number of STs involved, clonal transmission involved. Abbreviations: IQR- interquartile range; CIs – confidence intervals.

**Table S9: Epidemiological risk/protective factors for outbreak size for the subset of outbreak affecting at least 10 patients with descriptive summaries, univariable and multivariable associations with outbreak size (total number of patients infected/colonised per outbreak) ('10-or-more-patient' outbreaks model).** Descriptive summaries are provided considering the dataset of 140 outbreaks (out of 272 included in this review). Univariable associations are calculated from a dataset of outbreaks with complete data for each variable (shown in the 3rd column from the left). Multivariable associations are calculated from a dataset of outbreaks with complete data on that variable as well as all of the variables included in the final '10-or-more-patient' outbreaks model (publication type, number of outbreaks reported in study, transposon/IS involved in transmission, number of institutions, number of Enterobacterales STs involved, number of carbapenemase alleles involved, main Enterobacterales ST involved). Estimates and 95% confidence intervals (CIs) are reported on the original (non-$log_{10}$) scale, interpreted as relative changes in outbreak size (fold-change in size).

| Variable | Category | Number (%) missing | Number (%) outbreaks with data available | Median (IQR) or Number (%) | Range | Median (IQR) number patients affected | Univariable Fold-change (95% CIs) | p-value† | Multi-variable N | Multivariable‡ Fold-change (95% CIs) | p-value† |
|---|---|---|---|---|---|---|---|---|---|---|---|
| **Start year** | | 0 (0%) | 140 (100%) | 2016 (2013-2018) | 2010-2021 | - | 0.98 (0.94-1.02) | 0.311 | 133 | 0.99 (0.96-1.03) | 0.718 |
| **Continent** | | | | | | | | | | | |
| | **Europe** | 0 (0%) | 140 (100%) | 69 (49.3%) | - | 30 (19-49) | *ref* | 0.376 | 133 | *ref* | 0.547 |
| | **Africa** | - | - | 4 (2.9%) | - | 34 (21-60) | 1.14 (0.52-2.51) | 0.749 | 133 | 1.43 (0.75-2.73) | 0.272 |
| | **Asia** | - | - | 29 (20.7%) | - | 22 (14-47) | 0.77 (0.55-1.10) | 0.148 | 133 | 0.85 (0.62-1.16) | 0.290 |
| | **Australasia** | - | - | 11 (7.9%) | - | 44 (13-87) | 1.09 (0.63-1.89) | 0.741 | 133 | 0.98 (0.64-1.51) | 0.943 |
| | **North America** | - | - | 16 (11.4%) | - | 16 (13-22) | 0.67 (0.44-1.04) | 0.075 | 133 | 0.82 (0.53-1.26) | 0.355 |
| | **South America** | - | - | 11 (7.9%) | - | 26 (20-39) | 0.84 (0.49-1.44) | 0.526 | 133 | 0.83 (0.55-1.26) | 0.379 |
| **Country** | | | | | | | | | | | |
| | **China** | 0 (0%) | 140 (100%) | 18 (12.9%) | - | 27 (14-54) | *ref* | 0.713 | 133 | *ref* | 0.958 |
| | **Australia** | - | - | 11 (7.9%) | - | 44 (13-87) | 1.35 (0.71-2.56) | 0.349 | 133 | 1.12 (0.66-1.91) | 0.668 |
| | **Canada** | - | - | 1 (0.7%) | - | 65 (65-65) | 2.4 (0.49-11.91) | 0.280 | 133 | 1.53 (0.28-8.34) | 0.619 |
| | **Denmark** | - | - | 5 (3.6%) | - | 30 (25-72) | 1.27 (0.40-4.01) | 0.671 | 133 | 1.34 (0.38-4.74) | 0.651 |
| | **Italy** | - | - | 9 (6.4%) | - | 23 (20-48) | 1.19 (0.62-2.27) | 0.595 | 133 | 1.07 (0.63-1.83) | 0.798 |
| | **Other** | - | - | 43 (30.7%) | - | 26 (18-44) | 1.08 (0.69-1.67) | 0.734 | 133 | 1.03 (0.71-1.51) | 0.861 |
| | **Poland** | - | - | 13 (9.3%) | - | 27 (13-81) | 1.42 (0.72-2.78) | 0.302 | 133 | 1.60 (0.75-3.39) | 0.223 |
| | **Singapore** | - | - | 1 (0.7%) | - | 61 (61-61) | 2.26 (0.46-11.17) | 0.316 | 133 | 1.33 (0.29-6.11) | 0.714 |
| | **Spain** | - | - | 14 (10%) | - | 26 (14-44) | 1.06 (0.59-1.89) | 0.851 | 133 | 1.07 (0.67-1.70) | 0.783 |
| | **UK** | - | - | 10 (7.1%) | - | 36 (13-60) | 1.22 (0.65-2.31) | 0.530 | 133 | 0.87 (0.50-1.51) | 0.624 |
| | **USA** | - | - | 15 (10.7%) | - | 16 (12-20) | 0.76 (0.44-1.32) | 0.329 | 133 | 0.88 (0.52-1.50) | 0.637 |
| **Institution number** | | 3 (2.1%) | 137 (97.9%) | 1 (1-3) | 1-9 | - | 1.08 (1.02-1.15) | 0.005 | 133 | 1.10 (1.04-1.16) | **0.001** |
| **Institution type** | | | | | | | | | | | |
| | **Tertiary** | 7 (5%) | 133 (95%) | 81 (60.9%) | - | 23 (16-45) | *ref* | 0.434 | 128 | *ref* | 0.564 |
| | **Hospital (unspecified)** | - | - | 4 (3%) | - | 14 (12-31) | 0.71 (0.32-1.57) | 0.394 | 128 | 0.78 (0.41-1.48) | 0.448 |
| | **LTCF** | - | - | 2 (1.5%) | - | 64 (38-89) | 1.36 (0.45-4.14) | 0.585 | 128 | 2.02 (0.52-7.79) | 0.304 |
| | **Network** | - | - | 42 (31.6%) | - | 30 (14-70) | 1.29 (0.93-1.78) | 0.119 | 128 | 1.23 (0.88-1.71) | 0.229 |
| | **Secondary** | - | - | 4 (3%) | - | 28 (12-62) | 1.00 (0.45-2.19) | 0.995 | 128 | 0.99 (0.47-2.05) | 0.969 |

| Variable / Category | Number (%) missing | Number (%) outbreaks with data available | Median (IQR) or Number (%) | Range | Median (IQR) number patients affected | Univariable Fold-change (95% CIs) | p-value† | Multi-variable N | Multivariable‡ Fold-change (95% CIs) | p-value† |
|---|---|---|---|---|---|---|---|---|---|---|
| **Institution capacity** | 91 (65%) | 49 (35%) | 821 (600-1300) | 334-2420 | - | 1.00 (1.00-1.00) | 0.730 | 48 | 1.00 (1.00-1.00) | 0.222 |
| **Ward number** | 58 (41.4%) | 82 (58.6%) | 3 (2-6) | 1-9 | - | 1.04 (0.99-1.1) | 0.122 | 78 | 1.02 (0.97-1.07) | 0.394 |
| **Ward types*** | | | | | | | | | | |
| **Medical ICU** | 49 (35%) | 91 (65%) | 52 (57.1%) | - | 24 (15-50) | 1.09 (0.79-1.49) | 0.600 | 87 | 0.92 (0.70-1.21) | 0.545 |
| **General surgical** | - | - | 29 (31.9%) | - | 23 (17-44) | 0.95 (0.69-1.31) | 0.749 | 87 | 1.12 (0.85-1.49) | 0.419 |
| **Neurology** | - | - | 18 (19.8%) | - | 22 (14-58) | 0.88 (0.60-1.29) | 0.502 | 87 | 0.90 (0.64-1.27) | 0.553 |
| **Neonatal ICU** | - | - | 18 (19.8%) | - | 26 (19-42) | 0.94 (0.64-1.39) | 0.763 | 87 | 1.02 (0.74-1.42) | 0.894 |
| **General medical** | - | - | 20 (22%) | - | 26 (18-46) | 1.03 (0.70-1.51) | 0.867 | 87 | 1.03 (0.76-1.42) | 0.833 |
| **Surgical ICU** | - | - | 13 (14.3%) | - | 20 (13-47) | 0.90 (0.59-1.36) | 0.616 | 87 | 0.85 (0.58-1.27) | 0.430 |
| **Respiratory** | - | - | 13 (14.3%) | - | 40 (17-62) | 1.25 (0.81-1.93) | 0.313 | 87 | 1.13 (0.77-1.67) | 0.527 |
| **Paediatric** | - | - | 13 (14.3%) | - | 25 (19-62) | 1.07 (0.69-1.66) | 0.752 | 87 | 1.24 (0.85-1.82) | 0.258 |
| **Cardiology** | - | - | 12 (13.2%) | - | 48 (21-72) | 1.50 (0.95-2.37) | 0.083 | 87 | 1.50 (1.03-2.19) | **0.035** |
| **Emergency department** | - | - | 13 (14.3%) | - | 40 (22-61) | 1.28 (0.83-1.97) | 0.259 | 87 | 1.43 (0.94-2.19) | 0.095 |
| **Infectious disease department** | - | - | 7 (7.7%) | - | 23 (16-42) | 0.83 (0.50-1.38) | 0.456 | 87 | 0.94 (0.58-1.52) | 0.794 |
| **Haematology** | - | - | 9 (9.9%) | - | 41 (34-46) | 1.23 (0.74-2.04) | 0.424 | 87 | 1.36 (0.86-2.13) | 0.183 |
| **Rehabilitation** | - | - | 3 (3.3%) | - | 15 (14-19) | 0.56 (0.24-1.30) | 0.177 | 87 | 0.61 (0.28-1.33) | 0.208 |
| **Gastroenterology / Hepatology** | - | - | 9 (9.9%) | - | 19 (13-24) | 0.73 (0.43-1.24) | 0.244 | 87 | 0.92 (0.59-1.45) | 0.730 |
| **Orthopaedics** | - | - | 7 (7.7%) | - | 30 (18-50) | 1.01 (0.60-1.69) | 0.968 | 87 | 1.14 (0.69-1.91) | 0.600 |
| **Urology** | - | - | 10 (11%) | - | 46 (40-59) | 1.47 (0.89-2.42) | 0.132 | 87 | 1.63 (1.07-2.49) | **0.024** |
| **Transplant** | - | - | 3 (3.3%) | - | 21 (20-34) | 0.92 (0.39-2.14) | 0.836 | 87 | 0.75 (0.36-1.56) | 0.438 |
| **Endemicity pre-outbreak*** | 30 (21.4%) | 110 (78.6%) | 54 (49.1%) | - | 34 (18-49) | 1.24 (0.91-1.68) | 0.173 | 107 | 1.30 (1.00-1.69) | **0.050** |
| **Background screening*** | 87 (62.1%) | 53 (37.9%) | 46 (86.8%) | - | 24 (17-45) | 0.61 (0.32-1.16) | 0.128 | 50 | 0.41 (0.25-0.67) | **0.001** |
| **Outbreak Detection method** | | | | | | | | | | |
| **clinical** | 63 (45%) | 77 (55%) | 39 (50.6%) | - | 20 (14-44) | *ref* | 0.234 | 74 | *ref* | 0.935 |
| **retrospective** | - | - | 7 (9.1%) | - | 25 (12-51) | 0.95 (0.38-2.36) | 0.885 | 74 | 0.98 (0.37-2.60) | 0.965 |
| **screening** | - | - | 31 (40.3%) | - | 38 (19-72) | 1.40 (0.97-2.03) | 0.071 | 74 | 0.94 (0.66-1.33) | 0.717 |
| **Transmission mechanism*** | | | | | | | | | | |
| **Clonal** | 0 (0%) | 140 (100%) | 133 (95%) | - | 25 (14-48) | 0.86 (0.47-1.58) | 0.626 | 133 | 0.92 (0.54-1.54) | 0.739 |
| **Plasmid** | - | - | 62 (44.3%) | - | 32 (15-62) | 1.20 (0.92-1.57) | 0.171 | 133 | 1.04 (0.77-1.41) | 0.810 |
| **Transposon/IS** | - | - | 13 (9.3%) | - | 65 (27-100) | 1.94 (1.26-2.98) | **0.003** | 133 | 1.53 (1.03-2.26) | **0.035** |

| Variable Category | Number (%) missing | Number (%) outbreaks with data available | Median (IQR) or Number (%) | Range | Median (IQR) number patients affected | Univariable Fold-change (95% CIs) | p-value† | Multi-variable N | Multivariable‡ Fold-change (95% CIs) | p-value† |
|---|---|---|---|---|---|---|---|---|---|---|
| **Outbreak definition*** | | | | | | | | | | |
| **Epidemiological** | 60 (42.9%) | 80 (57.1%) | 19 (23.8%) | - | 20 (13-31) | 0.80 (0.54-1.19) | 0.262 | 77 | 0.83 (0.59-1.16) | 0.277 |
| **Clonal** | - | - | 48 (60%) | - | 19 (13-34) | 0.67 (0.48-0.93) | **0.019** | 77 | 0.62 (0.40-0.94) | **0.026** |
| **Plasmid** | - | - | 25 (31.2%) | - | 24 (15-41) | 1.11 (0.77-1.60) | 0.567 | 77 | 1.31 (0.90-1.89) | 0.150 |
| **Small MGE** | - | - | 3 (3.8%) | - | 20 (16-28) | 0.78 (0.24-2.50) | 0.600 | 77 | 0.57 (0.26-1.26) | 0.165 |
| **Gene-based** | - | - | 20 (25%) | - | 34 (19-66) | 1.63 (1.12-2.36) | **0.011** | 77 | 1.28 (0.87-1.89) | 0.201 |

* Compared to the absence of reporting of respective variables as the reference group.

†p-values are for a Wald test. p-values appearing in the reference ('ref') category row represent overall Wald-test p-values for that category, obtained using Satterthwaite's method for a type III ANOVA for mixed effects models. p-values <0.05 are shown in **bold**.

‡ Multivariate associations are adjusted for all 6 variables in the final core model: no outbreaks reported in study, publication year, number of institutions, start year, number of STs involved, clonal transmission involved.

Abbreviations: CIs – confidence intervals; IQR- interquartile range; IS – insertion sequence.

**Table S10: Genomic and microbiological risk/protective factors for outbreak size for the subset of outbreak affecting at least 10 patients with descriptive summaries, univariate and multivariable associations with outbreak size (total number of patients infected/colonised per outbreak) ('10-or-more-patient' outbreaks model).** Descriptive summaries are provided considering the dataset of 140 outbreaks (out of 272 included in this review). Univariable associations are calculated from a dataset of outbreaks with complete data for each variable (shown in the 3rd column from the left). Multivariable associations are calculated from a dataset of outbreaks with complete data on that variable as well as all of the variables included in the final '10-or-more-patient' outbreaks model (publication type, number of outbreaks reported in study, transposon/IS involved in transmission, number of institutions, number of Enterobacterales STs involved, number of carbapenemase alleles involved, main Enterobacterales ST involved). Estimates and 95% confidence intervals (CIs) are reported on the original (non-$log_{10}$) scale, interpreted as relative changes in outbreak size (fold-change in size).

| Variable | Category | Number (%) missing | Number (%) outbreaks with data available | Median (IQR) or number (%) | Range | Median (IQR) of number patients affected | Univariable Fold-change (95% CIs) | p-value† | Multi-variable N | Multivariable‡ Fold-change (95% CIs) | p-value† |
|---|---|---|---|---|---|---|---|---|---|---|---|
| **Number Enterobacterales spp.** | | 0 (0%) | 140 (100%) | 1 (1-4) | 1-4 | - | 1.09 (1.01-1.18) | **0.034** | 133 | 1.03 (0.89-1.19) | 0.667 |
| **Main Enterobacterales spp.** | | | | | | | | | | | |
| | ***K. pneumoniae* complex** | 0 (0%) | 140 (100%) | 87 (62.1%) | - | 28 (16-47) | *ref* | 0.171 | 133 | *ref* | 0.849 |
| | ***Citrobacter* spp.** | - | - | 4 (2.9%) | - | 38 (12-67) | 0.99 (0.46-2.14) | 0.979 | 133 | 0.63 (0.19-2.13) | 0.458 |
| | ***E. cloacae* complex** | - | - | 29 (20.7%) | - | 25 (12-47) | 0.88 (0.62-1.24) | 0.463 | 133 | 0.55 (0.07-4.24) | 0.566 |
| | ***E. coli*** | - | - | 8 (5.7%) | - | 52 (28-74) | 1.60 (0.92-2.79) | 0.098 | 133 | 0.99 (0.38-2.56) | 0.977 |
| | **Other** | - | - | 12 (8.6%) | - | 18 (14-24) | 0.68 (0.42-1.09) | 0.108 | 133 | 0.99 (0.19-5.03) | 0.988 |
| | **Number of Enterobacterales STs** | 0 (0%) | 140 (100%) | 1 (1-5.25) | 1-12 | - | 1.05 (1.02-1.09) | 0.002 | 133 | 1.04 (1.01-1.08) | 0.011 |
| **Main Enterobacterales STs*** | | | | | | | | | | | |
| | ***Other K. pneumoniae STs*** | 4 (2.9%) | 136 (97.1%) | 36 (26.5%) | - | 20 (14-31) | *ref* | **0.007** | 133 | - | **0.004** |
| | ***Citrobacter freundii STs*** | - | - | 3 (2.2%) | - | 65 (38-69) | 1.66 (0.69-3.97) | 0.251 | 133 | 1.25 (0.56-2.77) | 0.585 |
| | ***E. cloacae* complex STs** | - | - | 29 (21.3%) | - | 27 (12-48) | 1.28 (0.88-1.84) | 0.190 | 133 | 1.07 (0.76-1.50) | 0.698 |
| | ***E. coli STs*** | - | - | 7 (5.1%) | - | 66 (32-75) | 2.32 (1.28-4.18) | **0.006** | 133 | 2.13 (1.28-3.56) | **0.004** |
| | **ST11 *K. pneumoniae*** | - | - | 17 (12.5%) | - | 42 (21-49) | 1.46 (0.95-2.22) | 0.081 | 133 | 1.66 (1.13-2.42) | **0.010** |
| | **ST14 *K. pneumoniae*** | - | - | 8 (5.9%) | - | 62 (36-116) | 2.37 (1.35-4.15) | **0.003** | 133 | 1.82 (1.08-3.07) | **0.024** |
| | **ST15 *K. pneumoniae*** | - | - | 8 (5.9%) | - | 53 (23-76) | 1.73 (0.99-3.03) | 0.056 | 133 | 1.31 (0.81-2.13) | 0.273 |
| | **ST258 *K. pneumoniae*** | - | - | 10 (7.4%) | - | 20 (18-40) | 1.07 (0.64-1.79) | 0.803 | 133 | 1.01 (0.65-1.56) | 0.980 |
| | **ST307 *K. pneumoniae*** | - | - | 7 (5.1%) | - | 47 (36-61) | 2.11 (1.17-3.83) | **0.014** | 133 | 1.22 (0.72-2.08) | 0.458 |
| | **Other STs** | - | - | 11 (8.1%) | - | 19 (14-23) | 0.85 (0.51-1.39) | 0.509 | 133 | 0.72 (0.47-1.11) | 0.134 |
| **Non-Enterobacterales spp. present vs absent**** | | 0 (0%) | 140 (100%) | 2 (1.4%) | - | 28 (19-37) | 0.73 (0.24-2.17) | 0.565 | 133 | NA†† | NA†† |
| **Main Carbapenemase allele** | | | | | | | | | | | |
| | **NDM-1** | 0 (0%) | 140 (100%) | 26 (18.6%) | - | 26 (14-58) | *ref* | 0.056 | 133 | *ref* | 0.397 |
| | **Other IMP** | - | - | 3 (2.1%) | - | 18 (14-20) | 0.54 (0.22-1.32) | 0.174 | 133 | 0.74 (0.30-1.81) | 0.510 |
| | **IMP-4** | - | - | 5 (3.6%) | - | 96 (46-99) | 2.23 (1.06-4.68) | **0.034** | 133 | 1.48 (0.70-3.13) | 0.301 |
| | **Other KPC** | - | - | 4 (2.9%) | - | 16 (13-26) | 0.65 (0.29-1.49) | 0.307 | 133 | 0.93 (0.43-1.99) | 0.848 |
| | **KPC-2** | - | - | 29 (20.7%) | - | 26 (14-62) | 1.03 (0.69-1.55) | 0.876 | 133 | 0.86 (0.58-1.26) | 0.438 |

| Variable | Category | Number (%) missing | Number (%) outbreaks with data available | Median (IQR) or number (%) | Range | Median (IQR) of number patients affected | Univariable Fold-change (95% CIs) | p-value† | Multi-variable N | Multivariable‡ Fold-change (95% CIs) | p-value† |
|---|---|---|---|---|---|---|---|---|---|---|---|
| | **KPC-3** | - | - | 10 (7.1%) | - | 20 (15-24) | 0.67 (0.38-1.17) | 0.155 | 133 | 0.78 (0.46-1.31) | 0.343 |
| | **Mixed** | - | - | 13 (9.3%) | - | 41 (25-60) | 1.35 (0.81-2.23) | 0.245 | 133 | 0.86 (0.50-1.46) | 0.564 |
| | **Other NDM** | - | - | 9 (6.4%) | - | 19 (14-30) | 0.72 (0.41-1.27) | 0.256 | 133 | 0.80 (0.49-1.31) | 0.368 |
| | **OXA-48** | - | - | 18 (12.9%) | - | 38 (16-45) | 1.02 (0.64-1.61) | 0.932 | 133 | 1.20 (0.81-1.80) | 0.360 |
| | **Other OXA-48-like** | - | - | 10 (7.1%) | - | 47 (23-75) | 1.35 (0.78-2.33) | 0.287 | 133 | 1.41 (0.85-2.33) | 0.179 |
| | **Other VIM** | - | - | 4 (2.9%) | - | 13 (12-38) | 0.64 (0.29-1.44) | 0.278 | 133 | 0.80 (0.41-1.60) | 0.533 |
| | **VIM-1** | - | - | 9 (6.4%) | - | 20 (13-27) | 0.68 (0.38-1.24) | 0.209 | 133 | 0.82 (0.49-1.39) | 0.466 |
| **Main Carbapenemase gene family** | | | | | | | | | | | - |
| | **KPC** | 0 (0%) | 140 (100%) | 44 (31.4%) | - | 20 (14-47) | *ref* | 0.172 | 133 | *ref* | 0.179 |
| | **IMP** | - | - | 9 (6.4%) | - | 46 (22-96) | 1.54 (0.88-2.71) | 0.131 | 133 | 1.37 (0.78-2.40) | 0.270 |
| | **NDM** | - | - | 11 (7.9%) | - | 41 (28-52) | 1.51 (0.90-2.52) | 0.114 | 133 | 1.13 (0.81-1.59) | 0.459 |
| | **OXA-48** | - | - | 35 (25%) | - | 25 (14-46) | 1.04 (0.73-1.48) | 0.838 | 133 | 1.42 (0.99-2.05) | 0.058 |
| | **OXA-48-like** | - | - | 18 (12.9%) | - | 38 (16-45) | 1.15 (0.75-1.77) | 0.521 | 133 | 1.67 (1.04-2.69) | 0.036 |
| | **VIM** | - | - | 10 (7.1%) | - | 47 (23-75) | 1.50 (0.89-2.55) | 0.130 | 133 | 0.96 (0.59-1.58) | 0.887 |
| | **Mixed** | - | - | 13 (9.3%) | - | 14 (12-27) | 0.73 (0.43-1.23) | 0.232 | 133 | 1.01 (0.60-1.68) | 0.974 |
| **Number of carbapenemase alleles** | | 0 (0%) | 140 (100%) | 1 (1-1) | 1-2 | - | 1.73 (1.27-2.36) | **0.001** | 128 | 1.33 (0.96-1.83) | 0.083 |
| **Number of carbapenemase-assoc. plasmids** | | 19 (13.6%) | 121 (86.4%) | 1 (1-2) | 1-4 | - | 1.30 (1.13-1.51) | **<0.001** | 116 | 1.12 (0.96-1.31) | 0.146 |
| **Number of replicon types per isolate** | | 47 (33.6%) | 93 (66.4%) | 3 (2-5) | 1-10 | - | 1.06 (0.98-1.14) | 0.137 | 89 | 0.98 (0.91-1.05) | 0.560 |
| **Number of other MGEs per isolate** | | 61 (43.6%) | 79 (56.4%) | 1 (1-2) | 0-6 | - | 1.09 (0.98-1.22) | 0.114 | 77 | 1.06 (0.95-1.18) | 0.291 |
| **Other MGEs involved** | | | | | | | | | | | |
| **Any Integrons** | | 64 (45.7%) | 76 (54.3%) | 20 (26.3%) | - | 27 (17-46) | 1.02 (0.68-1.55) | 0.909 | 74 | 0.88 (0.60-1.31) | 0.540 |
| **Any Transposons** | | - | - | 50 (65.8%) | - | 32 (19-60) | 1.33 (0.91-1.93) | 0.138 | 74 | 1.21 (0.86-1.71) | 0.258 |
| | **Tn4401** | - | - | 19 (25%) | - | 28 (20-46) | 1.10 (0.73-1.65) | 0.648 | 74 | 1.04 (0.67-1.59) | 0.870 |
| | **Tn125-like** | - | - | 14 (18.4%) | - | 39 (25-58) | 1.21 (0.76-1.93) | 0.415 | 74 | 1.24 (0.81-1.91) | 0.321 |
| | **Tn3-like** | - | - | 8 (10.5%) | - | 38 (23-50) | 1.13 (0.64-2.00) | 0.661 | 74 | 0.98 (0.58-1.65) | 0.943 |
| | **Other Transposon** | - | - | 13 (17.1%) | - | 34 (19-61) | 1.18 (0.74-1.87) | 0.482 | 74 | 1.28 (0.83-1.98) | 0.263 |
| **Any Insertion Sequence** | | - | - | 26 (34.2%) | - | 32 (15-45) | 0.94 (0.65-1.37) | 0.757 | 74 | 1.17 (0.81-1.69) | 0.389 |
| | **IS26** | - | - | 8 (10.5%) | - | 34 (17-51) | 1.01 (0.57-1.79) | 0.964 | 74 | 1.01 (0.60-1.72) | 0.961 |
| | **Other IS** | - | - | 18 (23.7%) | - | 30 (15-44) | 0.92 (0.61-1.40) | 0.704 | 74 | 1.19 (0.81-1.75) | 0.381 |
| **Number of phenotypic resistances** | | 104 (74.3%) | 36 (25.7%) | 12 (10-14) | 4-19 | - | 0.96 (0.91-1.01) | 0.136 | 36 | 0.97 (0.93-1.00) ‡‡ | 0.079 |
| **Number of ARGs per isolate** | | 52 (37.1%) | 88 (62.9%) | 15 (10-19) | 3-25 | - | 1.01 (0.99-1.04) | 0.381 | 84 | 1.00 (0.98-1.03) | 0.863 |

| Variable | Category | Number (%) missing | Number (%) outbreaks with data available | Median (IQR) or number (%) | Range | Median (IQR) of number patients affected | Univariable Fold-change (95% CIs) | p-value† | Multi-variable N | Multivariable‡ Fold-change (95% CIs) | p-value† |
|---|---|---|---|---|---|---|---|---|---|---|---|
| **Number of VF genes per isolate** | | 93 (66.4%) | 47 (33.6%) | 3 (2-6.5) | 0-15 | - | 1.00 (0.95-1.06) | 0.890 | 46 | 1.00 (0.95-1.06) ‡‡ | 0.861 |
| **Main plasmid identity** | | | | | | | | | | | |
| | **pOXA-48**** | 29 (20.7%) | 111 (79.3%) | 8 (7.2%) | - | 41 (18-44) | 1.13 (0.65-1.98) | 0.658 | 107 | 1.26 (0.80-1.97) | 0.311 |
| **Main plasmid Inc type** | | | | | | | | | | | |
| | **IncFII** | 34 (24.3%) | 106 (75.7%) | 12 (11.3%) | - | 40 (14-64) | *ref* | 0.745 | 101 | *ref* | 0.593 |
| | **IncFIB** | - | - | 3 (2.8%) | - | 26 (18-51) | 0.82 (0.30-2.22) | 0.690 | 101 | 0.63 (0.25-1.56) | 0.313 |
| | **IncL** | - | - | 7 (6.6%) | - | 19 (14-38) | 0.60 (0.27-1.30) | 0.189 | 101 | 0.94 (0.54-1.66) | 0.842 |
| | **IncL/M** | - | - | 11 (10.4%) | - | 41 (26-44) | 1.31 (0.66-2.59) | 0.433 | 101 | 1.16 (0.68-1.98) | 0.572 |
| | **IncN** | - | - | 10 (9.4%) | - | 30 (16-40) | 0.89 (0.45-1.76) | 0.737 | 101 | 0.96 (0.55-1.66) | 0.879 |
| | **IncX** | - | - | 13 (12.3%) | - | 22 (15-45) | 0.82 (0.43-1.55) | 0.536 | 101 | 0.88 (0.53-1.46) | 0.625 |
| | **Other multireplicon** | - | - | 22 (20.8%) | - | 26 (15-63) | 0.93 (0.54-1.60) | 0.786 | 101 | 0.73 (0.48-1.13) | 0.156 |
| | **Other single replicon** | - | - | 28 (26.4%) | - | 29 (17-51) | 0.90 (0.52-1.56) | 0.707 | 101 | 0.78 (0.51-1.19) | 0.240 |
| **Main plasmid size (100kbp)** | | 55 (39.3%) | 85 (60.7%) | 0.70 (0.54-1.34) | 0.36-2.78 | - | 0.87 (0.69-1.11) | 0.258 | 83 | 0.79 (0.65-0.97) | **0.026** |
| **Number of ARGs on main plasmid** | | 81 (57.9%) | 59 (42.1%) | 5 (2-9) | 1-13 | - | 0.97 (0.92-1.02) | 0.202 | 56 | 0.94 (0.91-0.98) | **0.003** |
| **Number of small MGEs on main plasmid** | | 97 (69.3%) | 43 (30.7%) | 1 (1-2.5) | 1-10 | - | 0.91 (0.81-1.01) | 0.076 | 42 | 0.89 (0.81-0.98) | **0.014** |
| **Plasmid conjugation ability*** | | | | | | | | | | | |
| | **non-conjugative vs conjugative** | 102 (72.9%) | 38 (27.1%) | 7 (18.4%) | - | 26 (14-48) | 0.84 (0.44-1.59) | 0.580 | 38 | 0.98 (0.00-Inf) | 0.995 |

*The 'Other *K. pneumoniae* STs' category was the reference category for the Main ST variable, and comprised eight outbreaks with *K. pneumoniae* ST437, sever ST101, six ST51, five ST16, three each of ST17, ST405, ST78 and unspecified STs, two each of ST20 and ST54, and one each of ST1026, ST104, ST196, ST219, ST2253, ST23, ST231, ST2407, ST2497, ST25, ST26, ST283, ST2084, ST34, ST340, ST348, ST37, ST383, ST39, ST392, ST395, ST392, ST395, ST476, ST496, ST502, ST526, ST5551, ST571, ST661, ST716, ST76, ST859, ST873, ST968. The '*Escherichia coli* STs' category comprised five outbreaks with ST410, three ST313, 2 ST167, and one each of ST216, ST378, ST38, ST399, ST405, ST6388, and ST648, which were grouped together for statistical power. The 'Other STs' included three outbreaks with *Serratia marcescens* as the main Enterobacterales, two each of *Citrobacter farmerii* and *Klebsiella oxytoca* ST14, and one each of *Leclercia adenocarboxylata, Morganella morganii, Mixta calida, Proteus mirabilis, Salmonella* Goldcoast ST358, *Klebsiella variicola* ST15, *Klebsiella variicola* ST3456, *Klebsiella oxytoca* ST220, *Klebsiella oxytoca* ST29, and *Klebsiella aerogenes* ST233. The '*Enterobacter cloacae* complex STs' comprised six outbreaks ST89, four unspecified STs, three each of ST11, ST66, ST78, ST90, two of ST121, ST171, ST177, ST182, ST93, and one each of ST1015, ST1283, ST1303, ST131, ST134, ST14, ST190, ST198, ST269, ST310, ST428, ST595, ST69, ST873, ST94, and ST97. The '*Citrobacter freundii* STs' comprised three outbreaks ST18, two each of ST22 and ST65, and one each of ST523, ST98, and one unspecified ST.

** Compared to reference group of 'absent' other Enterobacterales spp., or non-pOXA-48 plasmids for Main plasmid identity associations are compared to the absence of reporting of respective variables as the reference group.

†p-values are for a Wald test. p-values appearing in the reference ('ref') category row represent overall Wald-test p-values for that category, obtained using Satterthwaite's method for a type III ANOVA for mixed effects models. p-values <0.05 are shown in **bold**.

‡ Multivariate associations are adjusted for all 6 variables in the final core model: no outbreaks reported in study, publication year, number of institutions, start year, number of STs involved, clonal transmission involved.

†† The variable 'Non-Enterobacterales spp. present' could not be added to the core model for outbreak size affecting at least 10 patients due to data sparsity (i.e. no outbreaks with non-Enterobacterales species present had non-missing data for all core model variables).

‡‡ The variables ‘’ and ‘’ were not adjusted for publication type from the core model, as these variables were only non-missing for journal articles among outbreaks affecting at least 10 patients (i.e. no conference abstracts or preprints were represented in this data subset). Abbreviations: ARG – antimicrobial resistance genes; CIs – confidence intervals; conj. – conjugative; IQR – interquartile range; IS – insertion sequence; ; kbp – kilobase-pair; MGE – mobile genetic element; ST – multi-locus sequence type; VF – virulence factor.

**Table S11: Infection control measures reported for the subset of outbreak affecting at least 10 patients with descriptive summaries, univariable and multivariable associations with outbreak size (total number of patients infected/colonised per outbreak) ('10-or-more-patient' outbreaks model).** Descriptive summaries are provided considering the dataset of 140 outbreaks (out of 272 included in this review). Univariable associations are calculated from a dataset of outbreaks with complete data for each variable (shown in the 3rd column from the left). Multivariable associations are calculated from a dataset of outbreaks with complete data on that variable as well as all of the variables included in the final '10-or-more-patient' outbreaks model (publication type, number of outbreaks reported in study, transposon/IS involved in transmission, number of institutions, number of Enterobacterales STs involved, number of carbapenemase alleles involved, main Enterobacterales ST involved). Estimates and 95% confidence intervals (CIs) are reported on the original (non-$log_{10}$) scale, interpreted as relative changes in outbreak size (fold-change in size).

| Variable* | Number (%) missing | Number (%) outbreaks with data available | Median (IQR) or number (%) | Range | Median (IQR) of number patients affected | Univariable Fold-change (95% CIs) | p-value† | Multi-variable N | Multivariable‡ Fold-change (95% CIs) | p-value† |
|---|---|---|---|---|---|---|---|---|---|---|
| **Any IPC measure reported** | 0 (0%) | 140 (100%) | 64 (45.7%) | - | 24 (17-47) | 1.08 (0.82-1.41) | 0.587 | 133 | 1.06 (0.83-1.36) | 0.649 |
| **Any Patient IPC measure** | 76 (54.3%) | 64 (45.7%) | 53 (82.8%) | - | 26 (18-47) | 1.25 (0.69-2.26) | 0.453 | 61 | 0.87 (0.51-1.47) | 0.592 |
| **Enhanced screening** | 76 (54.3%) | 64 (45.7%) | 23 (35.9%) | - | 40 (20-68) | 1.53 (1.02-2.29) | **0.039** | 61 | 1.45 (0.97-2.16) | 0.071 |
| **Patient Cohorting** | - | - | 19 (29.7%) | - | 41 (20-64) | 1.42 (0.93-2.16) | 0.101 | 61 | 1.09 (0.75-1.59) | 0.643 |
| **Contact precautions** | - | - | 33 (51.6%) | - | 36 (19-48) | 1.29 (0.87-1.91) | 0.203 | 61 | 1.20 (0.83-1.74) | 0.334 |
| **Screening during hosp.** | - | - | 30 (46.9%) | - | 30 (18-47) | 1.20 (0.80-1.80) | 0.380 | 61 | 1.01 (0.71-1.45) | 0.939 |
| **Contact tracing** | - | - | 15 (23.4%) | - | 40 (23-65) | 1.48 (0.93-2.37) | 0.095 | 61 | 1.31 (0.86-1.99) | 0.200 |
| **Patient - Other** | - | - | 19 (29.7%) | - | 40 (21-74) | 1.33 (0.87-2.02) | 0.185 | 61 | 1.36 (0.95-1.96) | 0.094 |
| **Any HCW IPC measure** | 76 (54.3%) | 64 (45.7%) | 47 (73.4%) | - | 25 (18-46) | 1.01 (0.63-1.63) | 0.966 | 61 | 0.87 (0.57-1.31) | 0.493 |
| **HCW Screening** | 76 (54.3%) | 64 (45.7%) | 11 (17.2%) | - | 45 (23-66) | 1.51 (0.90-2.53) | 0.113 | 61 | 1.91 (1.23-2.99) | **0.005** |
| **HCW Education** | - | - | 22 (34.4%) | - | 40 (19-61) | 1.29 (0.86-1.94) | 0.22 | 61 | 1.10 (0.75-1.60) | 0.625 |
| **HCW Enhanced hand hygiene** | - | - | 17 (26.6%) | - | 23 (18-47) | 1.02 (0.65-1.59) | 0.947 | 61 | 1.01 (0.68-1.50) | 0.945 |
| **HCW Cohorting** | - | - | 12 (18.8%) | - | 42 (22-62) | 1.52 (0.92-2.49) | 0.098 | 61 | 1.01 (0.63-1.63) | 0.958 |
| **Outbreak response team** | - | - | 10 (15.6%) | - | 19 (14-45) | 0.95 (0.52-1.74) | 0.871 | 61 | 1.09 (0.65-1.82) | 0.736 |
| **HCW - Other** | - | - | 30 (46.9%) | - | 24 (18-47) | 1.03 (0.69-1.54) | 0.883 | 61 | 0.98 (0.68-1.40) | 0.894 |
| **Any Environmental IPC measure** | 76 (54.3%) | 64 (45.7%) | 52 (81.2%) | - | 24 (17-62) | 1.23 (0.72-2.09) | 0.440 | 61 | 1.51 (1.00-2.28) | 0.051 |
| **Environmental Screening** | 76 (54.3%) | 64 (45.7%) | 38 (59.4%) | - | 24 (16-58) | 1.05 (0.70-1.57) | 0.821 | 61 | 1.52 (1.07-2.15) | **0.019** |
| **Environmental Cleaning** | - | - | 34 (53.1%) | - | 34 (19-60) | 1.18 (0.79-1.76) | 0.416 | 61 | 1.19 (0.83-1.70) | 0.343 |
| **Equipment Replacement** | - | - | 14 (21.9%) | - | 34 (18-45) | 1.01 (0.62-1.65) | 0.954 | 61 | 1.09 (0.68-1.76) | 0.717 |
| **Ward closure** | - | - | 8 (12.5%) | - | 56 (24-88) | 1.70 (0.95-3.04) | 0.072 | 61 | 1.64 (0.93-2.87) | 0.084 |
| **Environmental - Other** | - | - | 8 (12.5%) | - | 42 (34-65) | 1.51 (0.82-2.78) | 0.185 | 61 | 1.47 (0.90-2.42) | 0.124 |

* Compared to not reporting that specific control measure, among outbreaks reporting any control measures.

†p-values are for a Wald test. p-values appearing in the reference ('ref') category row represent overall Wald-test p-values for that category, obtained using Satterthwaite's method for a type III ANOVA for mixed effects models. p-values <0.05 are shown in **bold**.

‡ Multivariate associations are adjusted for all 6 variables in the final core model: no outbreaks reported in study, publication year, number of institutions, start year, number of STs involved, clonal transmission involved. Abbreviations: IQR- interquartile range; CIs – confidence intervals; HCW- healthcare worker; IPC – infection prevention and control.

**Table S12: Compliance with the Outbreak Reports and Intervention Studies of Nosocomial Infection (ORION) checklist(25, 26) items for 98 studies for which study quality was assessed.** (See also Figure 5).

| ORION Checklist item | Good compliance | Poor compliance | Not relevant / Not appropriate |
|---|---|---|---|
| **Title/ abstract** | | | |
| 1. Title/abstract | 70 (71%) | 28 (29%) | 0 (0%) |
| **Introduction** | | | |
| 2. Introduction: Background | 84 (86%) | 14 (14%) | 0 (0%) |
| 3. Introduction: Type of paper | 66 (67%) | 32 (33%) | 0 (0%) |
| 4. Introduction: Dates | 65 (66%) | 33 (34%) | 0 (0%) |
| 5. Introduction: Objectives | 79 (81%) | 19 (19%) | 0 (0%) |
| **Methods** | | | |
| 6. Methods: Design | 68 (69%) | 30 (31%) | 0 (0%) |
| 7. Methods: Participants | 30 (31%) | 68 (69%) | 0 (0%) |
| 8. Methods: Setting | 15 (15%) | 83 (85%) | 0 (0%) |
| 9. Methods: Interventions | 13 (13%) | 85 (87%) | 0 (0%) |
| 10. Methods: Culturing & typing | 81 (83%) | 17 (17%) | 0 (0%) |
| 11. Methods: Infection related outcomes | 33 (34%) | 65 (66%) | 0 (0%) |
| 12. Methods: Economic outcomes | 0 (0%) | 0 (0%) | 98 (100%) |
| 13. Methods: Potential threats to internal validity | 18 (18%) | 80 (82%) | 0 (0%) |
| 14. Methods: Sample size | 0 (0%) | 0 (0%) | 98 (100%) |
| 15. Methods: Statistical methods | 73 (74%) | 25 (26%) | 0 (0%) |
| **Results** | | | |
| 16. Results: Recruitment | 0 (0%) | 0 (0%) | 98 (100%) |
| 17. Results: Outcomes estimation | 60 (61%) | 38 (39%) | 0 (0%) |
| 18. Results: Ancillary analyses | 53 (54%) | 45 (46%) | 0 (0%) |
| 19. Results: Harms | 16 (16%) | 82 (84%) | 0 (0%) |
| **Discussion/ Conclusion** | | | |
| 20. Discussion: Interpretation | 49 (50%) | 49 (50%) | 0 (0%) |
| 21. Discussion: Generalisability | 64 (65%) | 34 (35%) | 0 (0%) |

**Supplementary Figure S1: Effect size of risk/protective factors for outbreaks affecting at least 10 patients a) included in the final core multivariable model (<10% missing data) and b) not included in the final core multivariable model (either not retained in the core model through backwards selection or >10% missing data), but added back into the core model individually to obtain an adjusted estimate**. In both **a)** and **b)**, each point estimate (circle) and CIs (grey bars) are adjusted for all variables in the core model shown in **a)**. Only statistically significant risk/protective factors (*p<0.05*) are shown in **b)**. The core model in **a)** is based on 133 outbreaks with complete data, while the number of outbreaks included in the adjusted models to obtain each estimate in **b)** varies and is shown for each variable in brackets after the variable name.

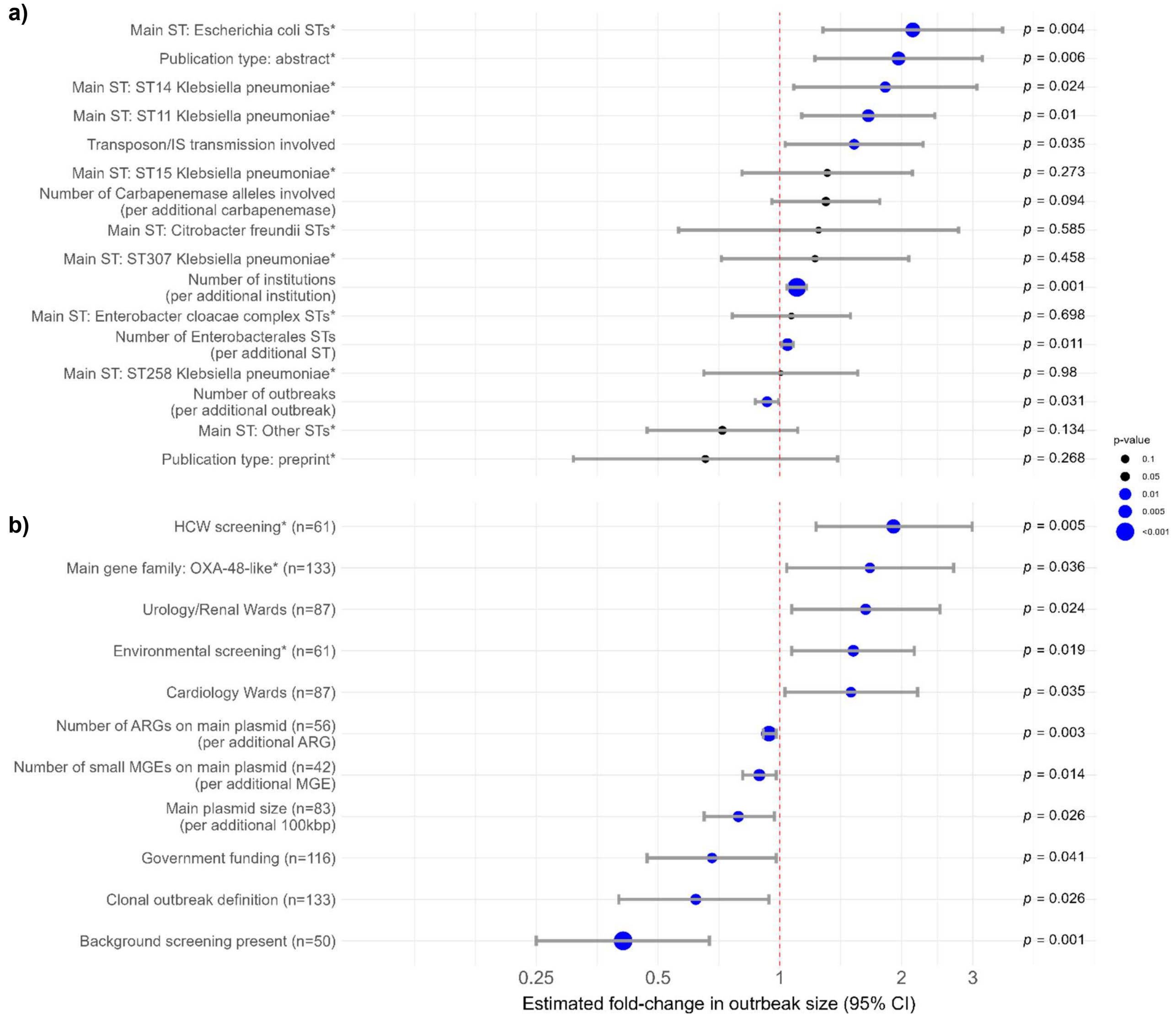

*The reference category used for publication type was peer-reviewed journal articles. The reference category for infection control measures was outbreaks reporting any infection control measures besides the given measure. The reference category used for main carbapenemase gene family was KPC, as it had the largest number of reports. The reference category for Enterobacterales ST was 'other *Klebsiella pneumoniae* STs' (STs besides ST11, ST14, ST15, ST307 or ST258). This 'other *K. pneumoniae* STs' category comprised eight outbreaks with *K. pneumoniae* ST437, sever ST101, six ST51, five ST16, three each of ST17, ST405, ST78 and unspecified STs, two each of ST20 and ST54, and one each of ST1026, ST104, ST196, ST219, ST2253, ST23, ST231, ST2407, ST2497, ST25, ST26, ST283, ST2084, ST34, ST340, ST348, ST37, ST383, ST39, ST392, ST395, ST392, ST395, ST476, ST496, ST502, ST526, ST5551, ST571, ST661, ST716, ST76, ST859, ST873, ST968. The '*Escherichia coli* STs' category comprised five outbreaks with ST410, three ST313, 2 ST167, and one each of ST216, ST378, ST38, ST399, ST405, ST6388, and ST648, which were grouped together for statistical power. The 'Other STs' included three outbreaks with *Serratia marcescens* as the main Enterobacterales, two each of *Citrobacter farmerii* and *Klebsiella oxytoca* ST14, and one each of *Leclercia adenocarboxylata, Morganella morganii, Mixta calida, Proteus mirabilis, Salmonella* Goldcoast ST358, *Klebsiella variicola* ST15, *Klebsiella variicola* ST3456, *Klebsiella oxytoca* ST220, *Klebsiella oxytoca* ST29, and *Klebsiella aerogenes* ST233. The '*Enterobacter cloacae* complex STs' comprised six outbreaks ST89, four unspecified STs, three each of ST11, ST66, ST78, ST90, two of ST121, ST171, ST177, ST182, ST93, and one each of ST1015, ST1283, ST1303, ST131, ST134, ST14, ST190, ST198, ST269, ST310, ST428, ST595, ST69, ST873, ST94, and ST97. The '*Citrobacter freundii* STs' comprised three outbreaks ST18, two each of ST22 and ST65, and one each of ST523, ST98, and one unspecified ST.
